# Quality and methodological heterogeneity of COVID-19 vaccine safety studies focusing on the myocarditis safety signal: A systematic review, meta-analysis and meta-regression

**DOI:** 10.1101/2025.06.18.25329834

**Authors:** Tu P. H. Trang, Marloes T. Bazelier, Olaf H. Klungel, Sophie H. Bots

## Abstract

**Background:** The risk of myocarditis following COVID-19 vaccines has been widely investigated, yielding highly variable effect estimates. It remains unclear how much of this variation can be explained by methodological differences and study quality. This study aimed to evaluate the methodology of observational studies on myocarditis risk post-COVID-19 vaccination and identify sources of heterogeneity.

**Methods:** We systematically searched PubMed and EMBASE for observational studies reporting relative risks of myocarditis after COVID-19 vaccination published between Dec 2022 and Oct 2023. Risk of Bias (RoB) was assessed using the ROBINS-I tool. Meta-analysis and multivariable meta-regression were conducted for studies addressing comparable population-intervention-comparator-outcome (PICO) questions.

**Results:** We included 30 studies, comprising 35 study designs. We identified considerable variability in design elements including risk window length (7-288 days), reference period timing (three different ways), and control for COVID-19 disease (five different approaches). Thirteen (37%) study designs had serious or critical overall RoB. We observed strongest heterogeneity between studies in general population for dose 2 of BNT162b2 and mRNA-1273 compared to unvaccinated individuals/reference period (I^2^=96%, prediction interval (PI) of relative risk 0.40 – 20.20; I^2^=92%, PI 1.23 – 88.63, respectively). Meta-regression for the former PICO indicated that after adjusting for age and sex, effect estimates of samples with 28-day risk window were 0.56 times (95% CI 0.43-0.72) lower than those with 7-day risk window. The effect of study design, outcome definition, approaches to handle COVID-19 infection and overall RoB were not significant in the meta-regression.

**Conclusions:** We observed considerable variation in design specifications including risk window length, reference period timing, and control for COVID-19 disease. Significant heterogeneity in effect estimates was identified, especially for second doses. Design choices like risk window length may explain some of this heterogeneity. Future vaccine safety studies should include sensitivity analyses to explore the effect of design choices on their findings.

## Background

The rapid development and distribution of Coronavirus disease 2019 (COVID-19) vaccines represents an unprecedented scientific accomplishment. The accelerated approval and massive roll-out of these vaccines also highlighted the importance of postmarketing safety monitoring to detect rare side effects for which the clinical trials were underpowered. Soon after vaccine roll-out, myocarditis was flagged as an important rare adverse event, and this signal was later confirmed by multiple real-world evidence studies across the world ^1,2^. However, the effect estimates varied considerably between studies. For example, the reported risk of myocarditis after the second dose of BNT-162b2 compared to no vaccination ranged between 0.79 and 29.7^3,4^. Such heterogeneous findings pose challenges to benefit-risk assessment for policy decision-making, as well as undermine public trust in vaccine safety.

The sources of heterogeneity in epidemiological studies can be roughly categorized into clinical diversity and methodological diversity^5^. In the context of vaccine safety, the clinical diversity entails the difference across studies in participant characteristics that affect their susceptibility to the adverse events, as well as the difference in vaccination policies and healthcare utilizations. Understanding these aspects helps to tailor the vaccination program to different populations and evaluate the transportability of evidence across different settings^6^. The methodological heterogeneity reflects the diversity in choice of designs and study parameters such as follow up length, selection of comparators, and covariate adjustments, as well as the degree of bias in each study ^5^.

Designing postmarketing vaccine safety study incurs special challenges, as vaccines are often administered by seasonal schedule which requires adjusting for time trends. In addition, vaccines are administered either to targeted populations or with high coverage, making it difficult to find appropriate control groups^7^. Various specialized methods have been developed and applied to accommodate these features, such as the case-centred analysis and self-controlled methods, yet their validity depends on meeting strict assumptions^7^. Prior simulation and empirical studies have suggested that the degree of biased results differs per study design, and that design parameters could partly explain the heterogeneity in study results ^6,8^. These findings may however have limited generalizability to the unique situation of the COVID-19 pandemic, with constraints in healthcare utilization and strong time trends in vaccination. It is thus of interest to characterise the methodologies of COVID-19 vaccine safety studies and their impact on the heterogeneity of effect estimates. These insights could be helpful in assessing the consistency and robustness of evidence and informing evidence synthesis and future study design selection in similar contexts.

Taking the potential risk of myocarditis following COVID-19 vaccination as a case study, we aimed to provide an overview of methodologies of inferential, comparative observational studies that evaluated COVID-19 vaccine safety signals. The first objective was to describe the study designs and analytical approaches used in observational studies on COVID-19 vaccine safety. The second objective was to assess the heterogeneity in findings across these studies and investigate the impact of study design and analysis choices on the observed heterogeneity.

## Methods

### Study design

We conducted a systematic review of the methodology of observational studies investigating the risk of myocarditis following COVID-19 vaccination. Effect estimates were grouped using the PICO (Population, Intervention, Comparator, Outcome) framework and meta-analysed within their PICO group to quantify statistical heterogeneity. Sources of heterogeneity were investigated using meta-regression. We followed the Preferred Reporting Items for Systematic Reviews and Meta-Analyses 2020 (PRISMA) checklist ^9^. The full study protocol is provided in Supplementary file 1.

### Eligibility criteria

We included studies fulfilling all following criteria:

1) Observational study that aimed to quantify the risk of myocarditis (with or without pericarditis) after COVID-19 vaccination;
2) The exposure was defined as at least one dose of any COVID-19 vaccine, where the exposed could either be an individual or a specific time period within an individual;
3) The comparator was defined as either no exposure to any COVID-19 vaccine or exposure to a different COVID-19 vaccination regimen, where the unexposed could either be an individual or a specific time period within an individual;
4) A comparative effect measurement (ratio or difference) was reported between the exposure and comparator groups;
5) Published in English as a peer-reviewed scientific article.
6) Published between December 11^th^, 2020 (first COVID-19 vaccine emergency use authorization) and October 9^th^, 2023^10^
7) Full-text available

Studies aimed at safety signal detection (which employ data mining techniques without prior hypothesis), or signal strengthening (such as disproportionality analysis, ecological studies, and crude before-after comparison) were excluded. Editorials, letters to editor, and commentaries were also excluded.

### Search strategy

Structured searches were conducted on PubMed and Embase on October 9^th^, 2023 to identify relevant records. The search strings combined keywords and subject heading terms related to COVID-19 vaccines, safety, and comparison of effect measure (Supplementary file 1).

### Study selection

The records were de-duplicated using Endnote X9^11^. Records were screened for inclusion by two independent reviewers (TT and SB) using ASReview, an open-source machine learning algorithm^12^. Discrepancies were resolved through discussion. The records were screened in two rounds with different machine-learning models to enhance the coverage of relevant records^13^, using the results of the first round as prior knowledge for the second round. Each screening round was stopped after 100 consecutive records were marked as “irrelevant”, or 50 consecutive irrelevant records after screening the first 1000 records.

After the title and abstract screening, the full text of the potentially eligible articles were assessed by three reviewers (TT, SB, MB). Each reviewer screened 33% and independently double checked 33% of the total records. Discrepancies between reviewers were resolved by discussion among the research team.

### Data extraction

Data extraction was performed by three reviewers (TT, SB, MB). Each reviewer extracted 33%, and one reviewer (TT) double checked randomly 50% of the total eligible studies. The following information was extracted from each study: study setting, data source and population; aspects of study design and statistical analysis, exposures, comparators, outcomes and effect estimates (Supplementary file 2). All primary analyses, secondary, sub-group and sensitivity analyses pertaining to the myocarditis outcome were extracted.

### Risk of bias assessment

The risk of bias (RoB) of each eligible study was assessed using the Risk of Bias in Non-randomized Studies of Interventions (ROBINS-I) tool ^14^. We adapted the tool to also cover assumptions specific to self-controlled and case control designs, as these were originally not included (Supplementary file 3). For self-controlled designs, we first checked whether the major validity assumptions as outlined by Whitaker et al (2006) were met^15^. Only self-controlled studies that fulfilled all validity assumptions underwent the ROBINS-I tool assessment. Each of three reviewers (TT, SB, MB) assessed 33% and independently double checked 33% of the total studies.

### Descriptive analysis

Continuous variables are presented as median and interquartile range. Categorical variables are summarised with frequencies and percentages. A post-hoc analysis was conducted to assess the consistency of results in studies where two study designs and two approaches to handle COVID-19 infection were used. Three agreement metrics were evaluated for each pair of effect estimates^16^: 1) Statistical significance agreement: whether two estimates and their confidence intervals are on the same side of the null; 2) Estimate agreement: whether the point estimate of one design/approach is within the 95% CI of the other design/approach; and 3) Standardized difference agreement: whether the absolute value of the standardized difference is < 1.96).

### Meta-analysis

Meta-analysis of primary analyses of the included studies were conducted to characterize the heterogeneity between studies. The effect estimates were grouped by PICO question: (P) General population or population with specific conditions, (I) Vaccine brand and dose, (C) unvaccinated individual/period or active comparator, and (O) outcome. As myocarditis was rare (<5% incidence), different ratio metrics (RR, OR, HR, IRR) could be combined^17^. Random-effect meta-analysis was conducted for PICOs with at least 3 studies. Tau-squared, I^2^ and prediction interval were calculated to measure degree of heterogeneity.

For studies that reported multiple effect estimates per PICO for overlapping samples (e.g. using different risk windows), a decision rule was applied to choose one single effect estimate per PICO per study^18^ (Supplementary file 1).

Univariate sub-group analyses were conducted for PICOs with at least three studies to explore the possible causes of heterogeneity in effect estimates between studies. The sub-groups included: 1) Age restriction of the study (general population, younger population only (<40 years old), or older population only (>50 years old); 2) Risk window length (within 14 days after vaccination, within 28 days after vaccination, or other lengths); 3) Study design (Cohort, self-controlled, or other designs); 4) Overall RoB (Low-moderate or serious-critical); 5) RoB of confounding domain (Low-moderate or serious-critical); 6) Outcome definition (myocarditis only, or myocarditis/pericarditis); and 7) Approach to handle COVID-19 infection in the analysis (No approach applied, approaches that measure direct effect of vaccination only (including censoring COVID-19 cases and excluding COVID-19 cases before and during the study), or other approaches).

Contour-enhanced funnel plots and Egger’s test of asymmetry were generated for PICOs with at least 10 studies to investigate small-study effects and differentiate asymmetry that is due to non-reporting biases from that due to other factors^19–22^.

### Examination of sources of heterogeneity by meta-regression

For PICOs with at least 10 studies without missing information on the explanatory variables and with considerable heterogeneity (*I*^2^ > 75%), multivariable random-effect meta-regression models were fitted to further investigate sources of heterogeneity, controlling for confounding effect of patient characteristics. The outcome was the log of the relative effect estimate. The explanatory variables included median age of the study sample, proportion of male in the study sample, study design, length of risk window, the approach to handle COVID-19 infection in the analysis, outcome definition, and overall RoB. All primary analyses, subgroup analyses and sensitivity analyses related to explanatory variables of interest were included in the model to increase statistical power and to estimate their between-study and within-study effects. Robust variance estimation method was employed to account for dependency between effect estimates from the same study, using the correlated and hierarchical effects working model^23^. The correlation between two effect sizes in the same study (p) is assumed to be constant and equal 0.7.

The base meta-regression model included two case-mix variables, age, and sex (Model 1). Next, the effect of methodological factors (study design, risk window length, outcome definition, handling of COVID-19 infection, and RoB) was investigated one by one, adjusted for age and sex (Model 2 to 6). For variables that varied both within a study and between studies (e.g. age and sex distribution, length of risk window, as studies often have subgroup/ sensitivity analysis regarding these variables), both the study means and the study-mean-centred values of these two variables were included in the model, to estimate separately their between-study and within-study effect, respectively^24,25^. The regression coefficients (β), their associated 95% CI and the Wald-style F test p-value of each variable (indicating the overall significance of a variable) were reported^25^.

## Results

Of all 10883 references identified, 266 full-text publications were reviewed. Thirty studies met the eligibility criteria and were included in systematic review (Supplementary figure S1)^3,4,26–53^. These studies comprised 35 designs (as five studies used two designs) and 36 distinct PICOs.

### General characteristics

The eligible studies were mostly conducted in Western Europe (14 studies, 47%) and North America (7 studies, 23%) (Table 1). Vaccination registries were the main data source for exposure ascertainment (20 studies, 67%), while the data sources for outcome and covariate ascertainment were more diverse. The two most common study designs for primary analysis were cohort (15 studies, 50%) and self-controlled designs (11 studies, 37%). Twenty-seven studies (87%) evaluated vaccine safety in the general population, while four (13%) targeted population with special conditions.

**Table 1.**
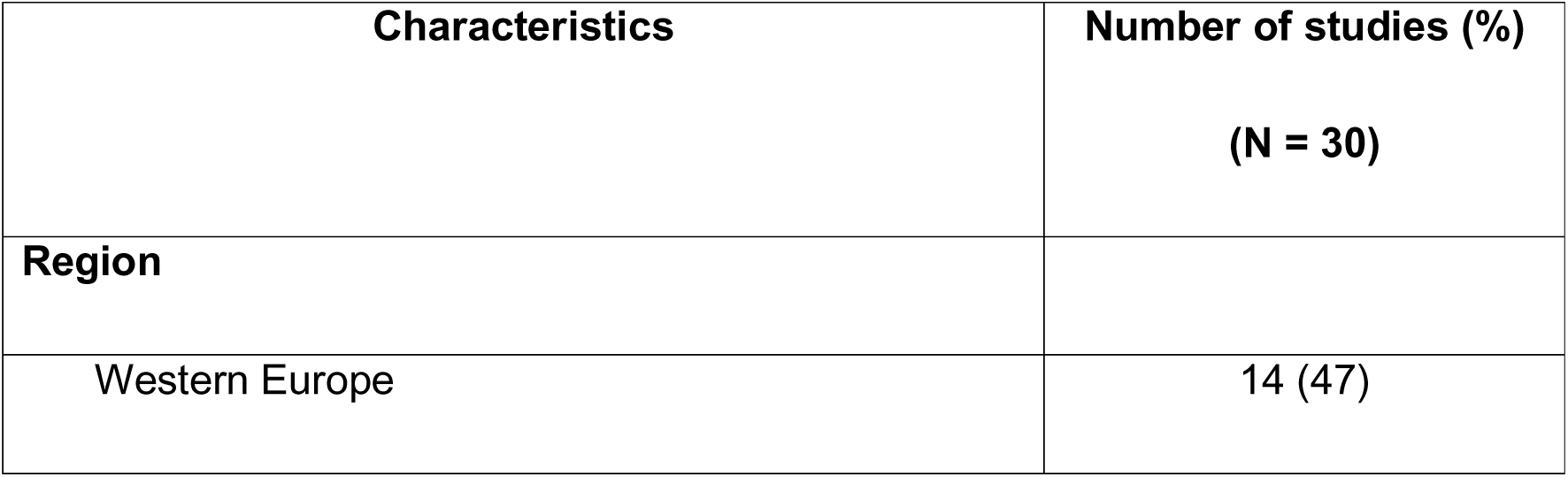
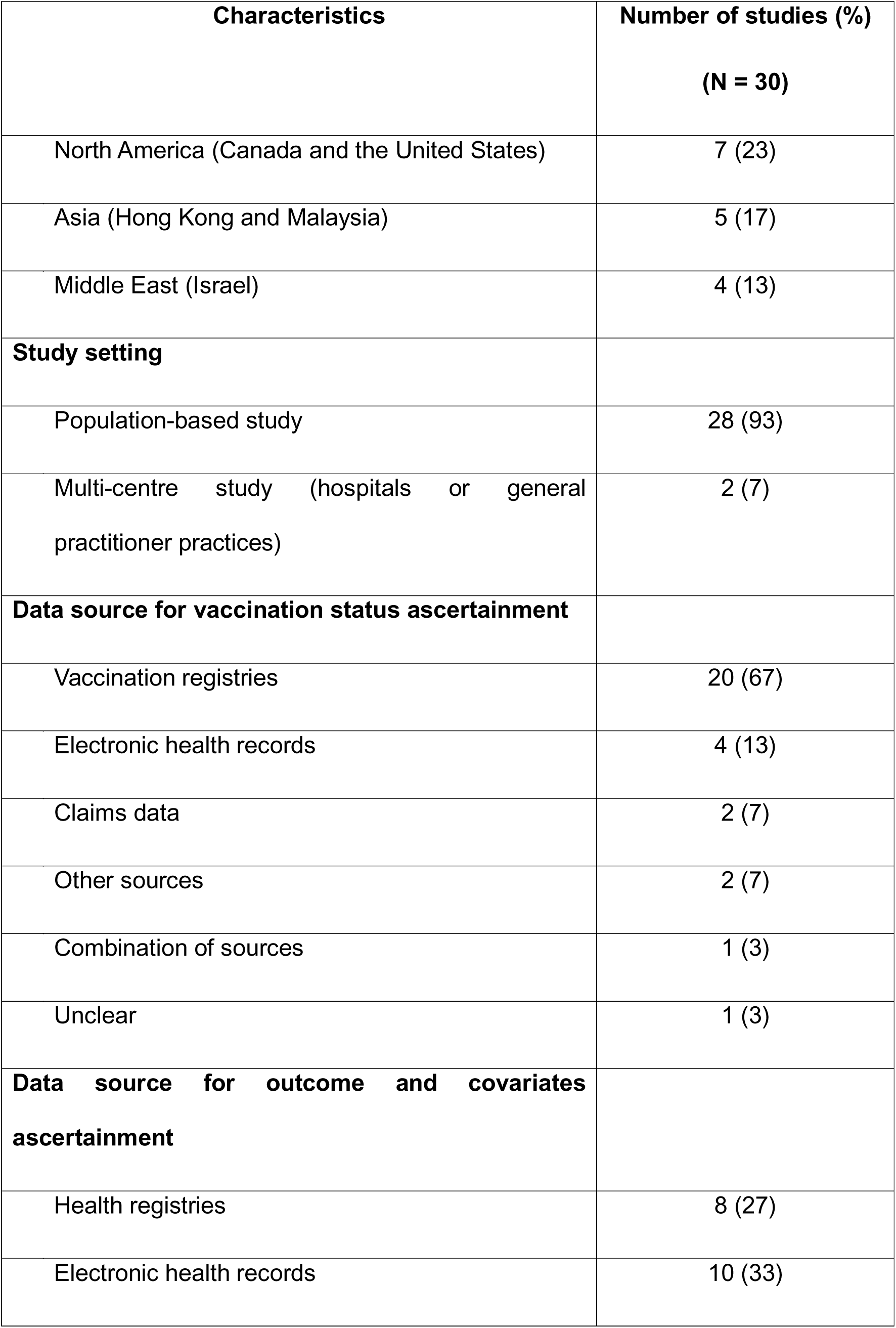
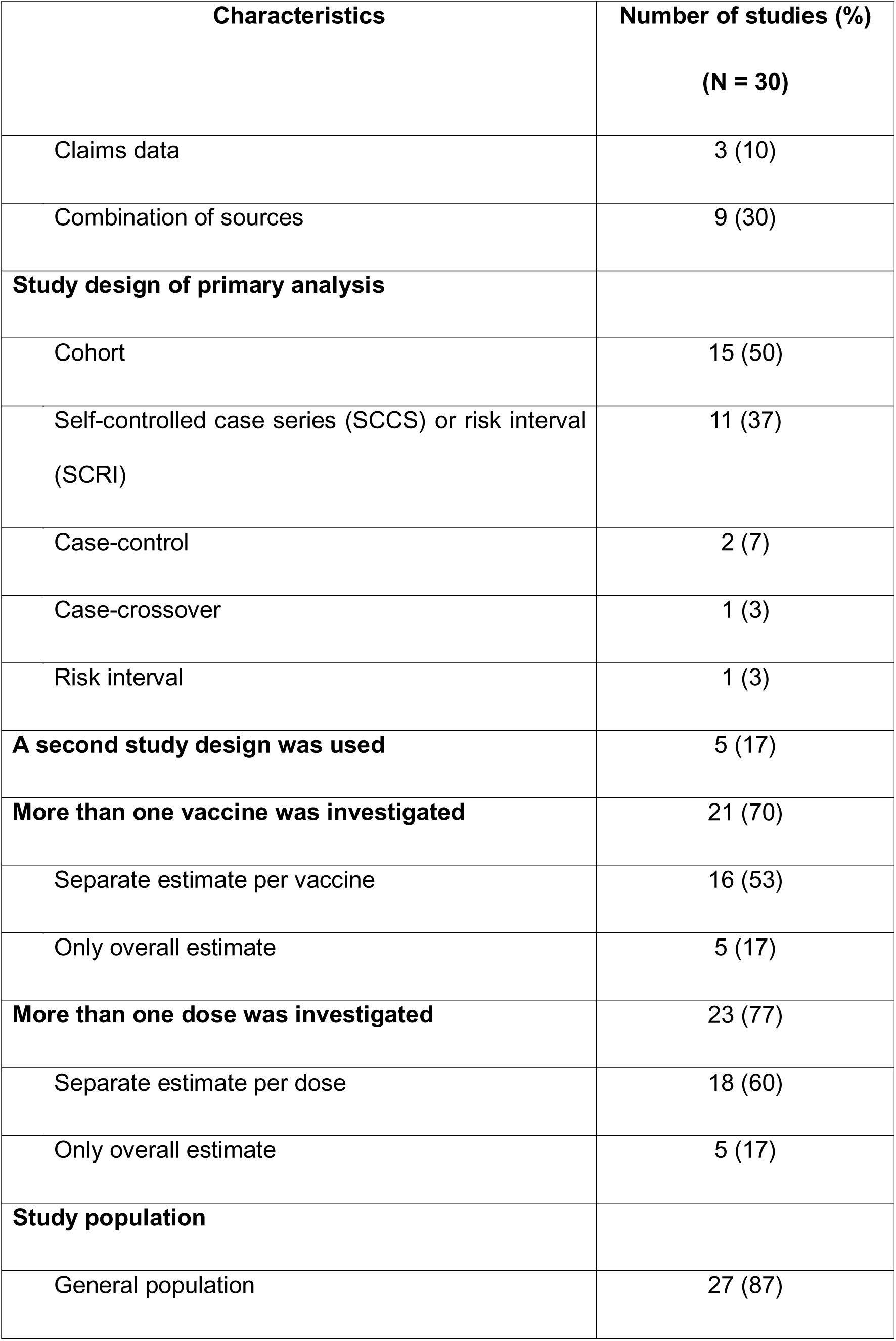
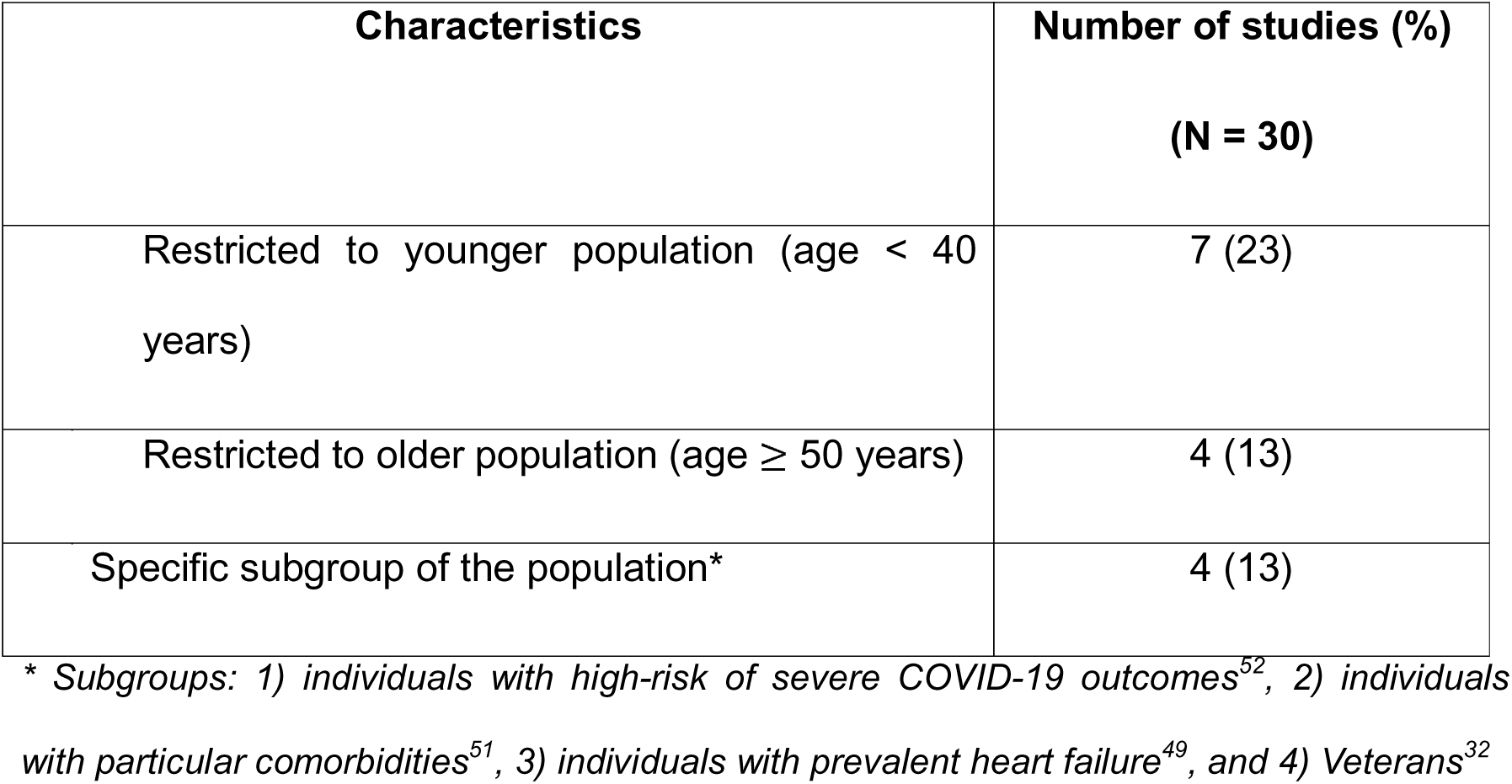
General characteristics of 30 eligible studies.

| Characteristics | Number of studies (%)<br>(N = 30) |
| --- | --- |
| **Region** |  |
| Western Europe | 14 (47) |

| **Characteristics** | **Number of studies (%)**<br>**(N = 30)** |
| --- | --- |
| North America (Canada and the United States) | 7 (23) |
| Asia (Hong Kong and Malaysia) | 5 (17) |
| Middle East (Israel) | 4 (13) |
| **Study setting** |  |
| Population-based study | 28 (93) |
| Multi-centre study (hospitals or general practitioner practices) | 2 (7) |
| **Data source for vaccination status ascertainment** |  |
| Vaccination registries | 20 (67) |
| Electronic health records | 4 (13) |
| Claims data | 2 (7) |
| Other sources | 2 (7) |
| Combination of sources | 1 (3) |
| Unclear | 1 (3) |
| **Data source for outcome and covariates ascertainment** |  |
| Health registries | 8 (27) |
| Electronic health records | 10 (33) |
| Claims data | 3 (10) |
| Combination of sources | 9 (30) |
| **Study design of primary analysis** |  |
| Cohort | 15 (50) |
| Self-controlled case series (SCCS) or risk interval (SCRI) | 11 (37) |
| Case-control | 2 (7) |
| Case-crossover | 1 (3) |
| Risk interval | 1 (3) |
| **A second study design was used** | 5 (17) |
| **More than one vaccine was investigated** | 21 (70) |
| Separate estimate per vaccine | 16 (53) |
| Only overall estimate | 5 (17) |
| **More than one dose was investigated** | 23 (77) |
| Separate estimate per dose | 18 (60) |
| Only overall estimate | 5 (17) |
| **Study population** |  |
| General population | 27 (87) |
| Restricted to younger population (age < 40 years) | 7 (23) |
| Restricted to older population (age ≥ 50 years) | 4 (13) |
| Specific subgroup of the population* | 4 (13) |

\* Subgroups: 1) individuals with high-risk of severe COVID-19 outcomes<sup>52</sup>, 2) individuals with particular comorbidities<sup>51</sup>, 3) individuals with prevalent heart failure<sup>49</sup>, and 4) Veterans<sup>32</sup>

Among 36 distinct PICOs investigated in the included studies (Supplementary Table S1), most PICOs were on a specific dose of a specific vaccine (23 PICOs), with unvaccinated individuals or reference period as comparator (29 PICOs). Seven PICOs had active comparators, either comparing mRNA and BNT-162b2^26,32,34,35,42^ or comparing heterologous and homologous vaccination courses^28^. The two PICOs examined by the most studies were effect of first dose and effect of second dose of BNT-162b2 compared to unvaccinated individual/period (14 studies per PICO). When a combination of doses was studied, it could be defined as “first and second dose”^29,32^, “first or second dose”^34,36,46^ or “any dose”^31,33,38,39,43^.

### Diversity of study design parameters

#### Definition of “unvaccinated individuals” and “reference period” as comparator

There were five cohort studies that used “unvaccinated individuals” as comparator. One used historical controls^49^, two matched vaccinated and unvaccinated on the vaccination date^4,29^, and the other two assigned an artificial index date to unvaccinated controls^41,43^.

For two case-control studies, one used hospital controls^39^, while one selected individuals without the outcome in the whole population^53^.

Reference period within an individual was used as comparator in five cohort studies and all self-controlled studies (total 18 studies, 60%). Among them, ten studies selected all periods before, between and after risk period as reference period^3,27,31,33,40,44–46,50,52^, six studies only used period before first exposure^30,36–38,47,51^, and two only use period after the risk period^34,48^.

#### Outcome definition and ascertainment

Fifteen studies (50%) included only myocarditis as outcome, eleven studies (37%) included myocarditis and pericarditis as a composite outcome, and four studies (13%) included both definitions^27,33,34,39^.

#### Risk window length

There were 15 studies (50%) that used at least two different risk window lengths in their analysis. The most chosen risk window length was 28-day post-vaccination (15 studies, 50%), followed by 7-day post-vaccination (12 studies, 40%).

Day 0 (day of vaccination) was included in the risk window in 13 studies (43%)^3,4,29,31,32,34,36–38,50–52^ and excluded in 7 studies (23%)^28,30,45,47–49,53^. Two studies included both risk windows with and without day 0,^27,40^ and three studies took day 0 as a separate risk window^33,44,46^. The risk window definition was unclear in five studies^35,39,41–43^.

#### Control for confounding

In all 35 study designs, age and sex were controlled for. Seasonality, comorbidities, outcome history and socioeconomic status were controlled for in 83%, 80%, 77% and 69% designs, respectively.

COVID-19 infection could act as both confounder and mediator of the association between vaccination and myocarditis. In three out of 35 designs (11%), cases with COVID-19 infection before the study period were excluded^34,35,48^, and two designs excluded COVID-19 cases both before and during study period^33^. COVID-19 cases were censored in 7 designs (20%)^28,29,32,36,37,51^. In two designs (6%), COVID-19 infection was included as covariate in the model^42,53^. In 20 designs (57%), COVID-19 infection was not considered in the main analysis, among them 6 designs had sensitivity analysis that used one of the above approaches^3,27,30,40,44,50^.

### Risk of bias in the included studies

Overall risk of bias was low for one (3%), moderate for 21 (60%), serious for 10 (29%), and critical for 3 (9%) designs (Figure 1). Most of this variability could be attributed to the confounding domain, where 31% of designs were rated as either critical or serious risk of bias. All 13 included self-controlled studies met the major validity assumptions, and most (11 studies) implemented correct adjustments to ensure that exposure was not dependent on occurrence of the outcome. The full RoB assessment for each study is presented in Supplementary file 4.

**Figure 1.**
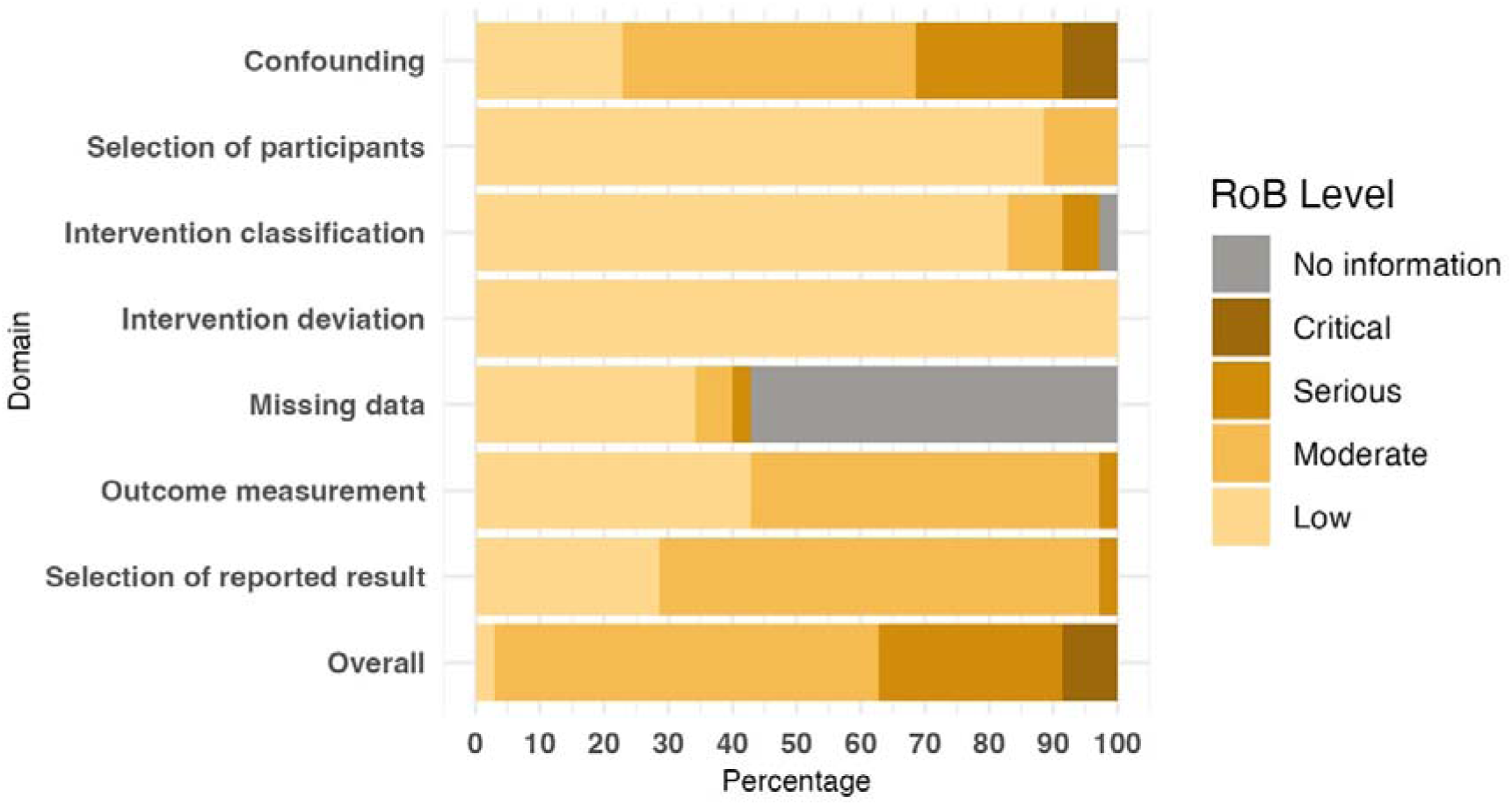
Proportion of risk of bias levels among 35 designs

### Studies that used two study designs

There were four studies that used two study designs to answer the exact same PICO questions, resulting in a total of 31 pairs of effect estimates. Three studies^3,27,36^ conducted SCCS and cohort, and one^33^ conducted SCCS and case-control. Generally, the effect estimates from SCCS were higher than other designs (Figure 2). In 7 pairs (23%) one of the three agreement metrics was not met.

**Figure 2.**
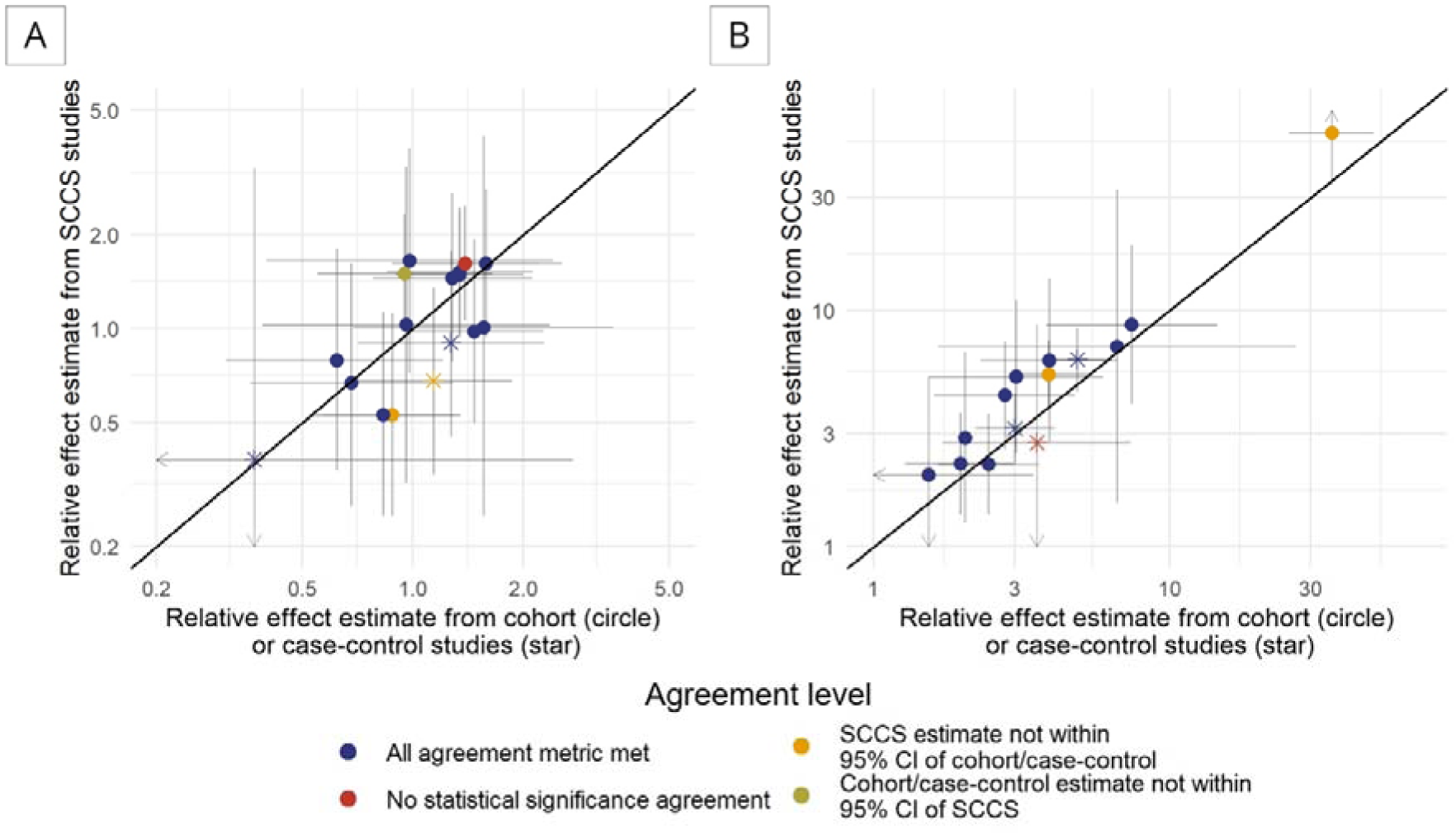
Comparison of pairs of effect estimates in studies that used two study designs to answer the exact same PICO questions. A) Pairs where both relative effect estimates were lower or equal to 2. B) Pairs where one of the relative effect estimates was higher than 2. Arrowheads indicate that the bounds of the confidence interval exceed the limit of the plot.

### Studies that used two approaches to handle COVID-19 infection

There were 10 studies that applied two approaches to control for COVID-19 infection in the exact same PICO questions, resulting in a total of 40 pairs of effect estimates ^3,27,32,36,38,40,44,45,48,50^. Most pairs met all agreement metrics; there were only two pairs (5%) without statistical significance agreement (Figure 3).

**Figure 3.**
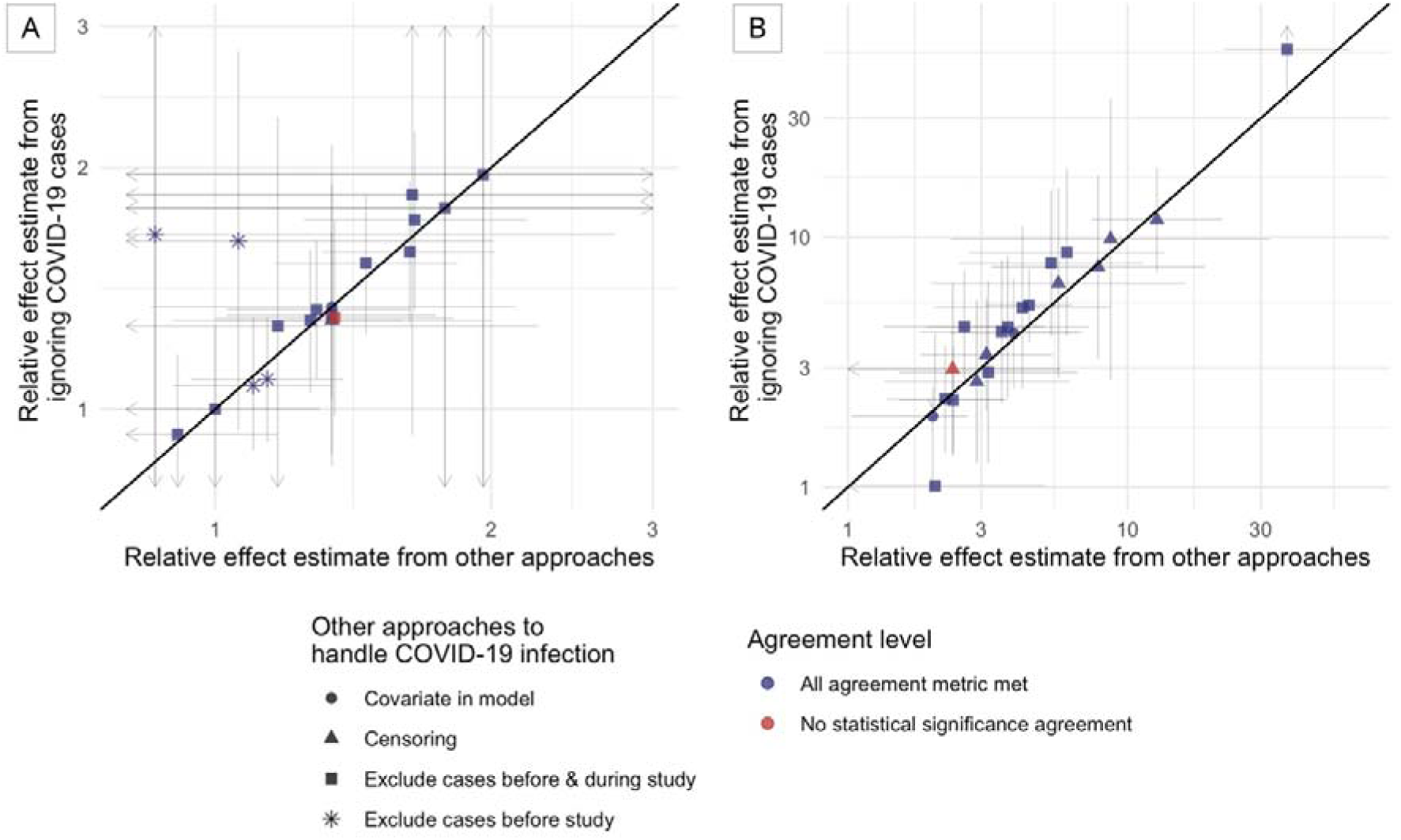
Comparison of pairs of effect estimates in study that used two approaches to handle COVID-19 infection in an exact same PICO question. A) Pairs where both relative effect estimates were lower or equal to 2. B) Pairs where one of the relative effect estimates was higher than 2. Arrowheads indicate that the bounds of the confidence interval exceed the limit of the plot.

### Meta-analysis

We performed a meta-analysis for 12 PICOs answered in more than two studies (Supplementary Table S2). The funnel plots suggested low risk of publication bias (Supplementary Figure S2-S3). Heterogeneity was highest between estimates for the second dose of BNT-162b2 versus unvaccinated individuals/person-time in the general population (15 studies, I^2^ = 95.6% (95% CI: 91.4 – 96.8)) and lowest between estimates for the second dose of CoronaVac versus unvaccinated individuals/person-time in the general population (3 studies, I^2^ = 0% (95% CI 0-89.6)). For the first and second doses of both BNT-162b2 and mRNA-1273 versus unvaccinated individuals/person-time in the general population, we observed larger heterogeneity for the second dose compared to the first dose (indicated by higher I^2^ and wider prediction interval), even though the estimates came from the same set of studies (Figure 4).

**Figure 4.**
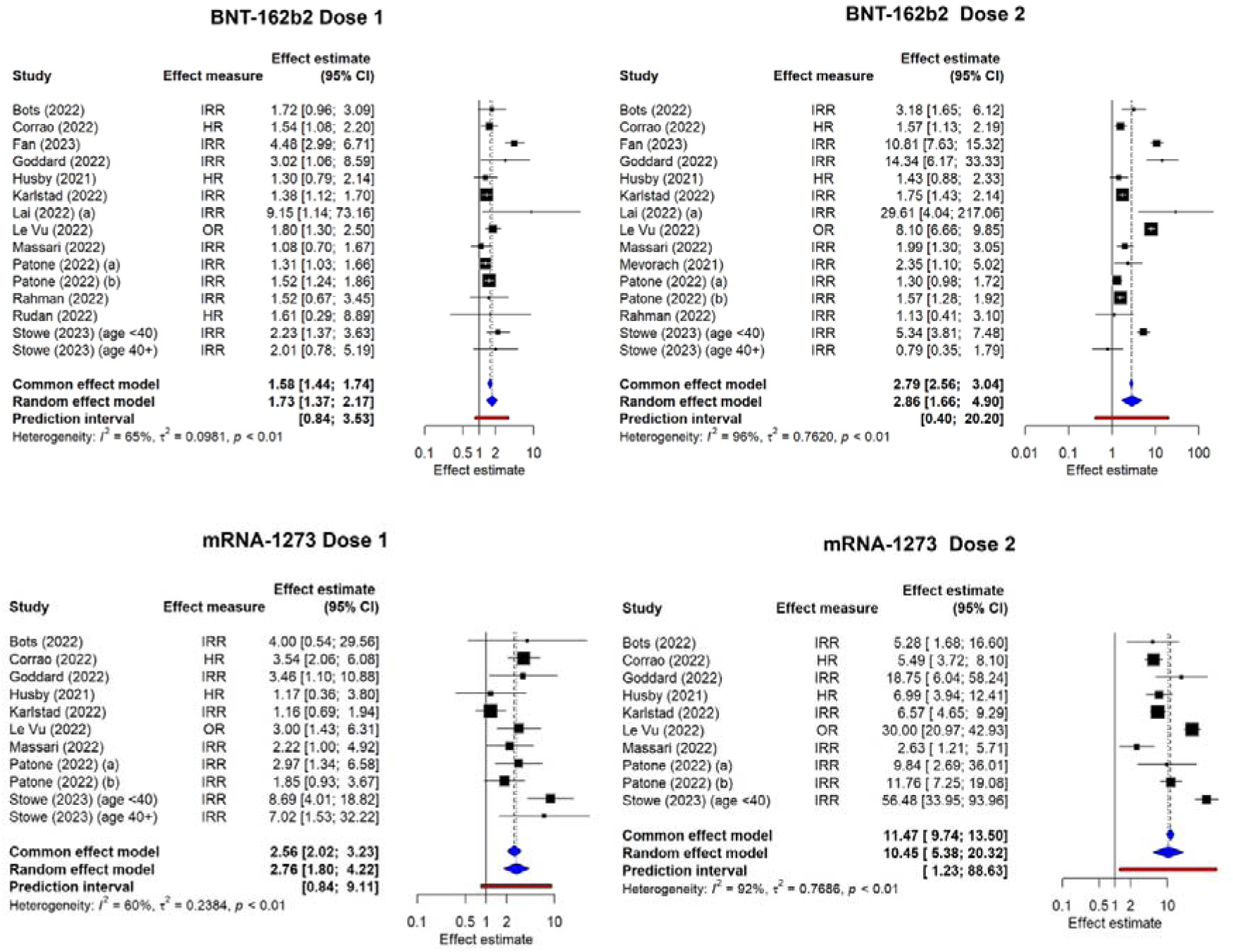
Meta-analysis of the risk of myocarditis following BNT-162b2 dose 1 (top left) and dose 2 (top right), and mRNA-1273 dose 1 (bottom left) and dose 2 (bottom right) compared with unvaccinated individuals/ reference periods, in general population. Note that the objective of the meta-analysis was to measure heterogeneity and not to estimate the pooled effect.

Subgroup analysis showed no clear effect of study design, outcome definition, or risk of bias (overall or confounding only) on heterogeneity between studies (Supplementary Tables S3-S9). Subgroup analyses were also conducted for age (12 PICOs), risk window length (9 PICOs), and approaches to handling COVID-19 infection (10 PICOs). These suggested effect estimates were higher in younger populations compared to the general or older population (statistically significant in 4/12 PICOs), and in studies using shorter risk windows compared to longer ones (statistically significant in 5/9 PICOs). There was no clear trend in the effect of approaches used to handle COVID-19 infection.

### Meta-regression

Meta-regression was performed for the only PICO with at least 10 studies: Dose 2 of BNT-162b2 compared with unvaccinated individuals/ reference period in general population (Table 2). The within-study estimates showed that compared to female-only populations, the average effect estimate in male-only study populations was 1.34 times greater (95% CI 1.07 – 1.67). For age, the average effect estimates were 0.55 times (95% CI:0.33-0.94) and 0.33 times (95% CI 0.14-0.79) lower in populations with a median age of 30-49 years and >49 years, respectively, compared to populations with a median age under 30 years. For risk window length, the average effect estimates were 0.56 times (0.42-0.72) lower in studies with a 28-day risk window compared to those with a 7-day risk window.

**Table 2.**
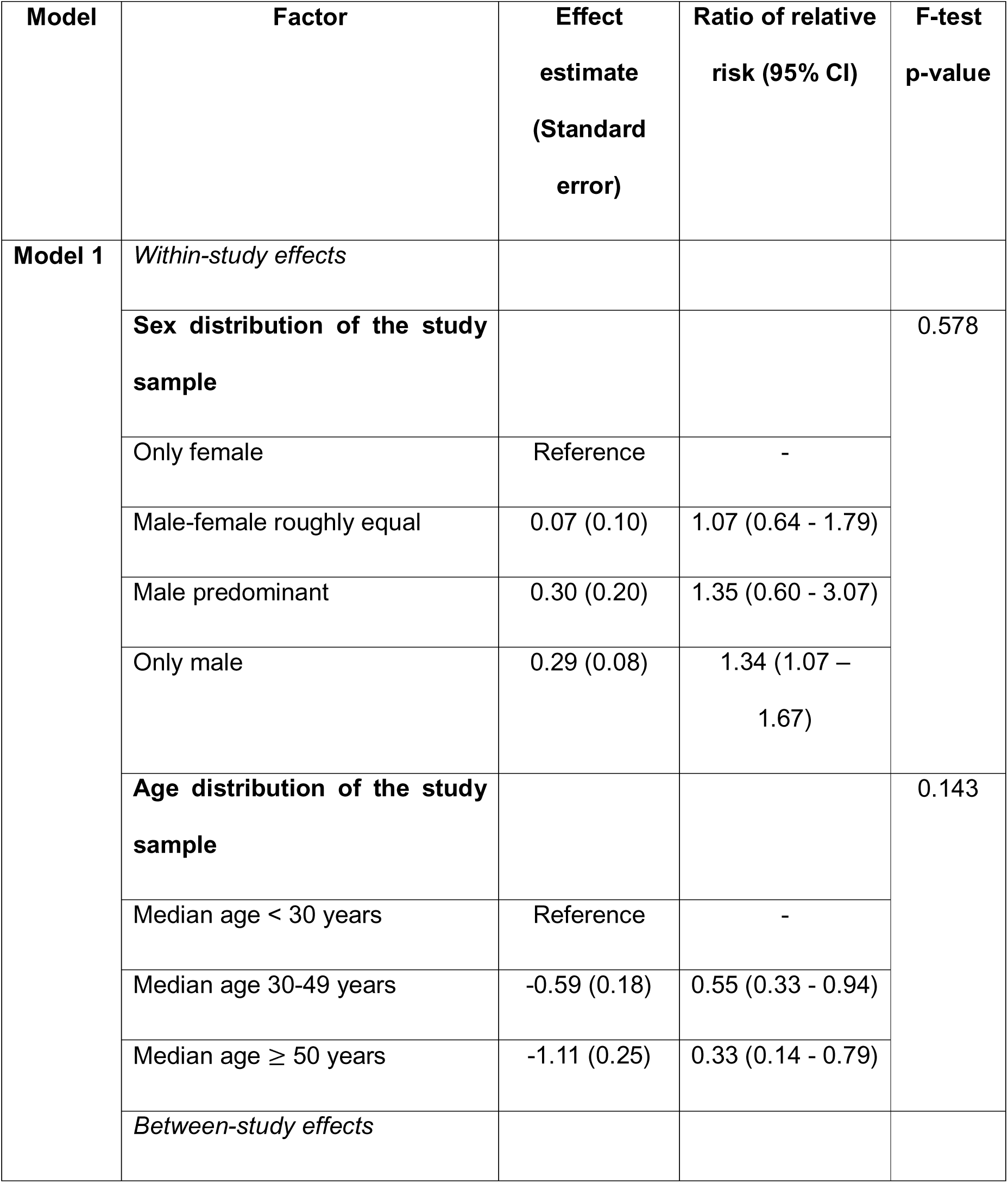
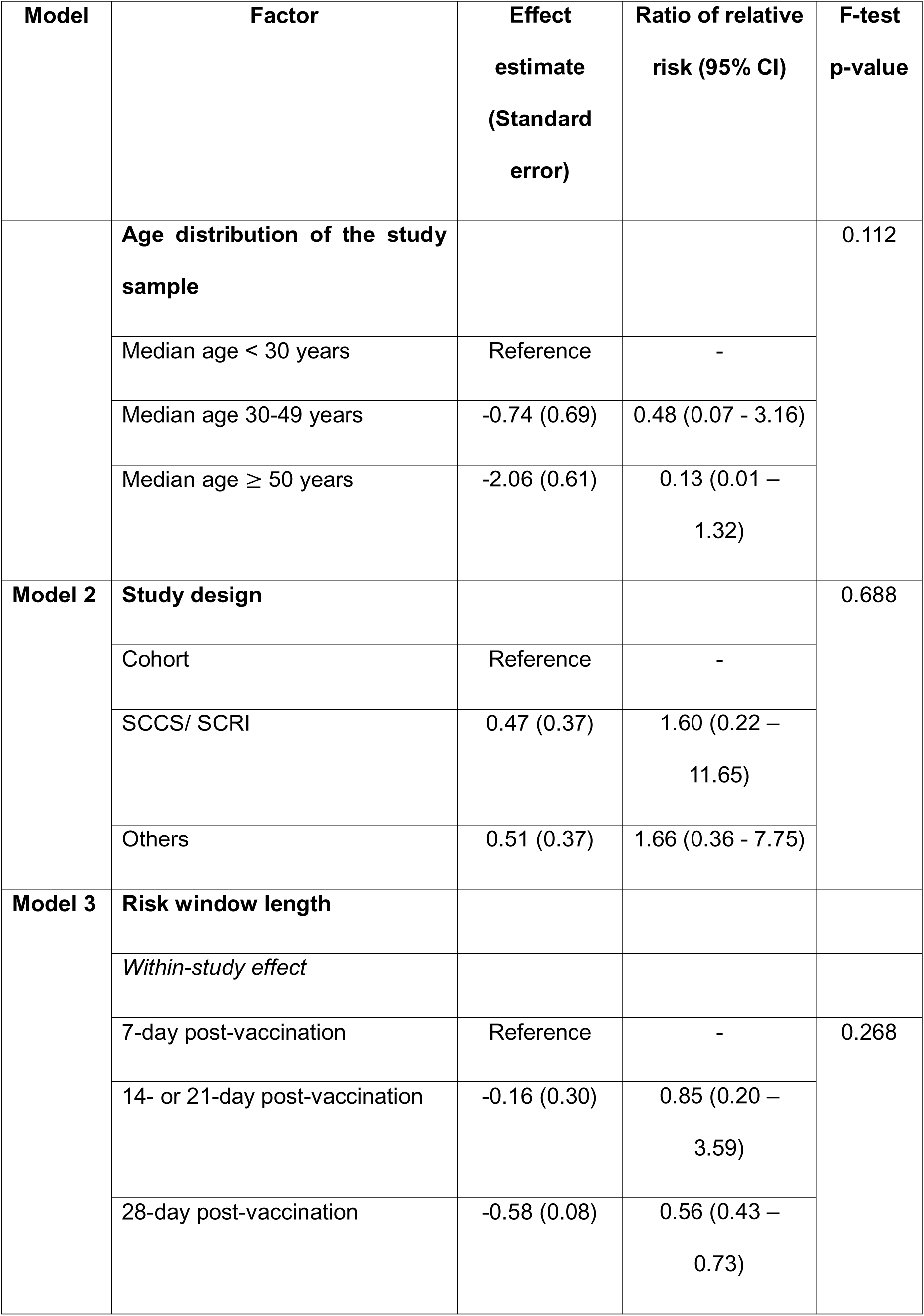
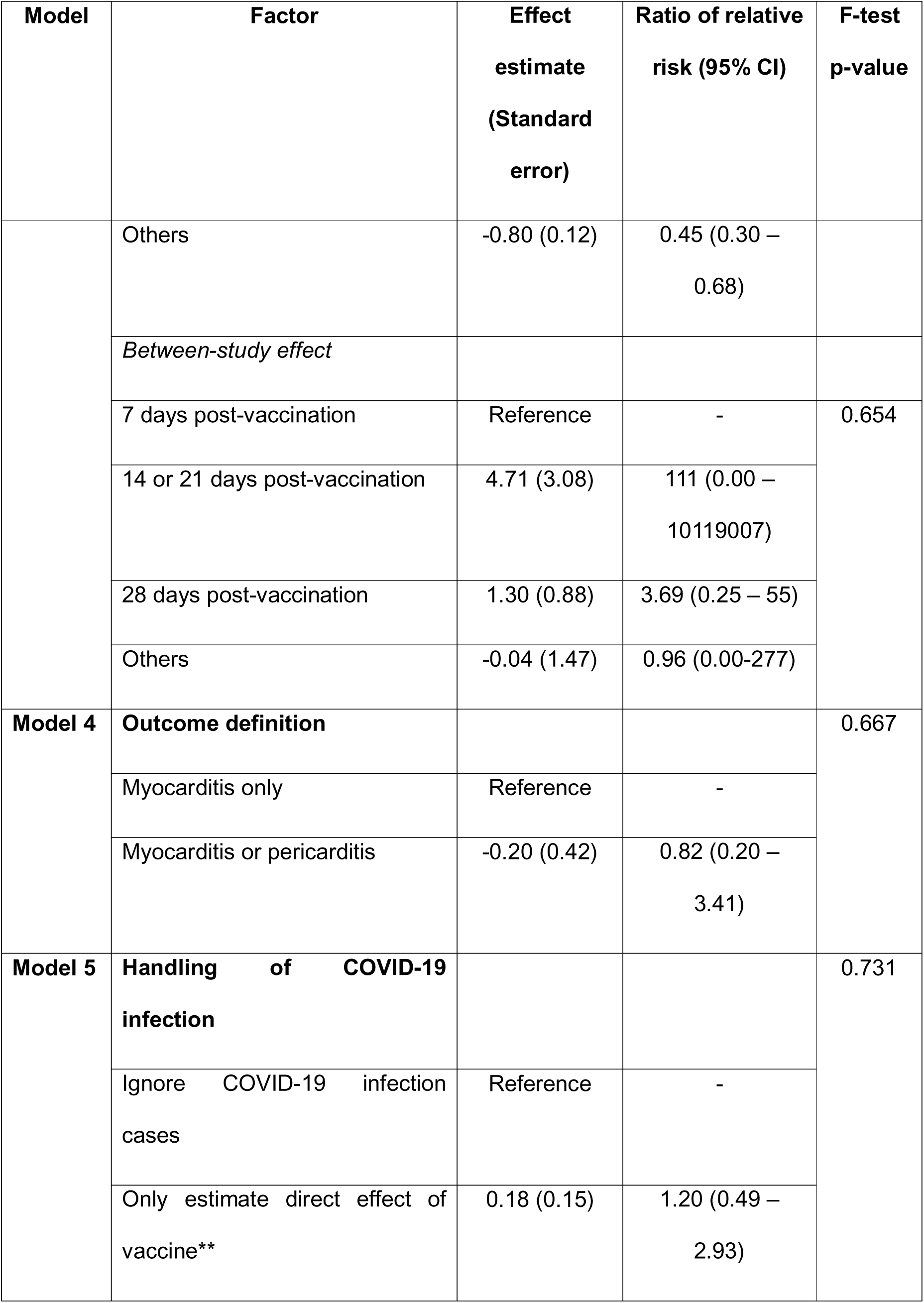
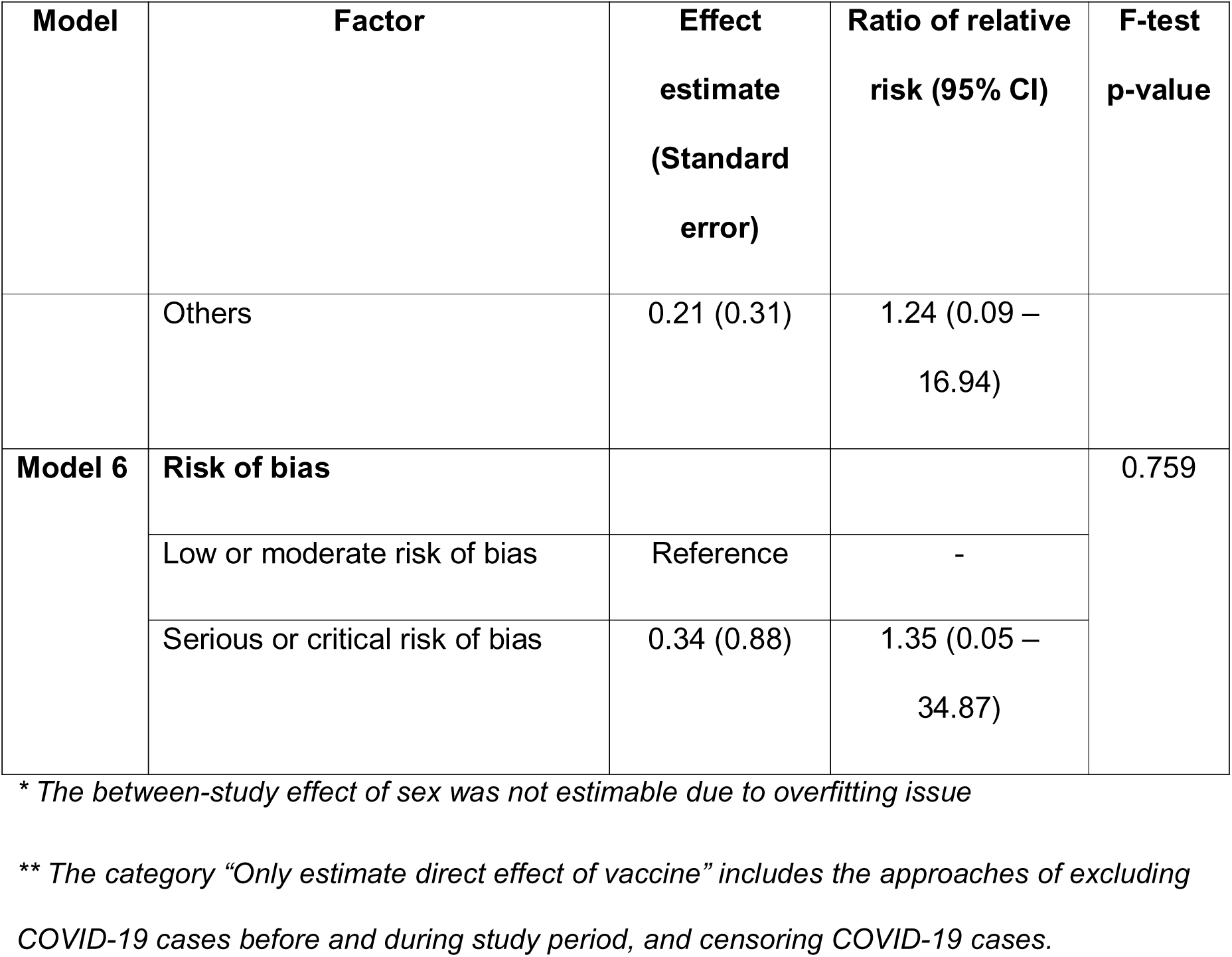
Results of meta-regression models for PICO of Dose 2 of BNT-162b2 compared with unvaccinated individuals/ reference period in general population. Model 1 includes effect of age and sex, model 2 to 6 include methodological factors one by one, adjusting for age and sex.

We were unable to estimate the between-study effect for sex due to overfitting and the between-study effect for age and risk-window length were not significant. The overall effects of sex, age, and risk window length were also not significant when tested using F-test (Table 2).

## Discussion

We systematically reviewed the methodology of 30 observational studies assessing the risk of myocarditis following COVID-19 vaccination, and employed meta-analysis and meta-regression to examine heterogeneity in effect estimates and the influence of design specifications. Substantial variability was observed in design choices, including risk window length, reference period timing, and approaches to account for COVID-19 infection. Approximately one-third of the studies were rated as having serious or critical risk of bias, primarily due to confounding and bias in outcome measurement. The highest heterogeneity in the effect estimates was observed for second doses of BNT-162b2 and mRNA-1273 compared to unvaccinated individuals/reference period in general population. Meta-regression suggested that longer risk windows were associated with smaller effect estimates, however its overall effect lacked significance.

In recent years there has been increasing adoption of the self-controlled designs, specifically SCCS and SCRI, to evaluate vaccine safety^54^. Our findings are consistent with this trend, in which SCCS was the second most common design, after the cohort design. The SCCS/SCRI overcomes several typical challenges in vaccine safety studies. First, only cases are included, thereby avoiding the challenges of selecting unvaccinated controls particularly relevant in high-vaccine-coverage settings like those for COVID-19 vaccines. Second, the SCCS/SCRI inherently controls for all time-invariant confounders, mitigating risks associated with unmeasured confounding often encountered in between-person designs^55^. The SCCS/SCRI also minimizes selection bias and exposure misclassification bias as only vaccinated individuals are studied^55^. There are, however, certain caveats with this design. Only relative risk estimation is available for this design, and it is susceptible to measurement error related to timing of outcome and exposure, and subsequently the definition of risk and control periods. Furthermore, validity of this design requires fulfilment of several exposure and outcome criteria, or adaptation of the design^15,56^. Encouragingly, most SCCS studies in our review adhered to these assumptions, a notable improvement compared to earlier systematic reviews^57,58^.

No single study design is universally optimal for all vaccine-outcome pairs. The selection of a suitable design should align with the characteristics of the outcome, risk of confounding and misclassification bias, and availability of data and resources^7,55^. Besides, it is also sensible to triangulate findings with more than one appropriate design. The agreement of results from different design could strengthen the evidence, and disagreement could shed light on the sources of bias^59^. Four studies in our review employed both SCCS and cohort or case-control designs and they mostly yielded consistent conclusions. It is important to note, however, that estimates from different designs reflect different estimands and need not be numerically identical. A prior simulation study also reported that the SCCS estimates were within 3% of a cohort gold standard, while a recent empirical evaluation study suggested that SCCS was less biased than cohort or case-control^8,60^.

A noteworthy finding was the higher heterogeneity in the effect estimates for second doses of BNT-162b2 and mRNA-1273 compared to their first doses. The first and second dose of each vaccine were investigated by the same set of studies, meaning that the set of source populations, study designs and other design specification were similar between the two PICOs. This may denote that the factors specifically pertain to second dose could play a role in explaining the effect heterogeneity. One factor is the interval between first and second dose, ranging from 21 days to up to 84 days in different countries^36,61–63^. Studies have shown that longer inter-dose intervals could significantly reduce the risk of post-vaccination myocarditis^64,65^. However, we could not investigate this factor in our analysis due to limited reporting in original papers. Another factor could be the increased level of residual (time-varying) confounding, as the second dose risk window was further away from the control window before vaccination, and individuals who chose to receive second dose could be more different from those who did not get vaccinated. Researchers should therefore pay great attention to the risk of time-varying and selection bias when designing a multi-dose vaccine safety study^55^.

Selecting an appropriate risk window is critical for unbiased effect estimation, particularly in SCCS/SCRI studies. If the chosen risk window is much longer or shorter or is placed so that it does not cover the true risk period, the relative incidence estimates could be biased toward the null^15,66^. The significant effects of several risk window categories on the magnitude of effect estimates identified in our meta-regression reaffirms the importance of careful consideration in defining risk window. Yet it is challenging to select the optimal risk window as clinical trials often provide little or no insight on the timing of specific adverse events. Vaccine safety studies, including those in our review, have often used pre-specified risk window based on literature, biological plausibility, expert opinions or convention^67^. It is recommended to include several (contiguous) periods, especially when there is uncertainty about the true risk period, albeit with potential increases in type I error^15^. Several data-driven methods to identify the optimal risk window have also been proposed, yet their adoption remains limited^66,68,69^.

The included studies demonstrated broad variation in how they accounted for COVID-19 infection cases in their analyses: exclude or censor cases with COVID-19 infection, include COVID-19 infection as a covariate in the model, or ignore the cases altogether. COVID-19 infection represents a time-varying confounder that is affected by past exposure when considering the COVID-19 vaccine – myocarditis relationship, as COVID-19 infection also increased the risk of myocarditis, and the risk of having an infection could be different between the control window and the risk window^45,70^. Thus, both self-controlled studies and cohort and case-control studies, especially those comparing different time periods, are susceptible to this confounding bias. A quantitative bias analysis of COVID-19 vaccine-myocarditis relationship in SCRI design showed that a strong association between the time-varying confounder and the outcome would lead to overestimation of the true effect if the confounder was ignored^70^. While our findings suggest that different approaches to handle COVID-19 infection may not substantially alter effect estimates, additional research is needed to investigate their implications further.

While our meta-regression models identified significant effects of certain levels of sex, age and risk window length, none of their overall effects was significant. These results should be interpreted cautiously considering the inherent drawbacks of meta-regressions. Due to the low number of studies in the meta-regression, the model may be underpowered to detect effects and be susceptible to confounding as only a limited number of variables could be included in the model^5^. Furthermore, with the lack of individual participant data, aggregate-data meta-regression is prone to ecological fallacy. We have tried to minimize this bias by using all available subgroup and sensitivity analyses and separating between-study and within-study effect (which is free from ecological bias) of certain variables^71^. However, the variability and incompleteness of reporting certain information across studies, such as age and sex distribution, hampered the granularity of our analysis. We thus call for a more comprehensive and uniform reporting of the participant and design characteristics in future studies, by using reporting checklist or core participant characteristics sets^72^. As suggested by Wang and colleagues (2022), researchers could also conduct sensitivity analyses over a designed matrix of alternative design specifications, with justification of the variations used in sensitivity analyses^6^. These practices would be particularly beneficial in evidence synthesis, enabling the standardization of results prior to meta-analysis to reduce heterogeneity, and the investigation of the sources of heterogeneity with higher precision and accuracy^72^.

Besides the limitations of the meta-regression method, the scarcity of data limited our ability to investigate the role of other factors such as data sources, outcome ascertainment methods, and types of reference period. Prior research has suggested data differences could play a role in heterogeneity of real-world evidence studyies^6^. Additionally, there is currently no official quality assessment tool for self-controlled studies. We have adapted the ROBINS-I tool to accommodate the features of self-controlled designs, however this adapted version has not been validated. Developing standardized reporting guidelines and quality assessment frameworks tailored to these designs could address these gaps.

## Conclusions

Our review identified substantial variability in design aspects such as risk window length, reference period timing, and accounting for COVID-19 disease, in studies investigating the myocarditis safety signal of COVID-19 vaccines. Considerable heterogeneity in effect estimates was observed, particularly for second doses. Design choices like risk window length may explain at least some of this heterogeneity. Future vaccine safety studies should include thorough sensitivity analyses to explore the effect of design and analytical choices on their findings, which could in turn strengthen the usefulness of real-world evidence in vaccine safety evaluation, informing evidence synthesis and benefit-risk assessments of vaccines.

## Data Availability

All data produced in the present work are contained in the manuscript

https://github.com/trangtph/Myocarditis_Vaccine_SR_public

## Supplementary material

Supplementary file 1: Study protocol

Supplementary file 2: Data extraction form

Supplementary file 3: Adapted ROBINS-I tool for SCCS/SCRI studies

Supplementary file 4: ROBINS-I assessment results

Supplementary Figure S1: PRISMA low diagram

Supplementary Figure S2. Contour-enhanced funnel plot for Dose 2 of BNT-162b2 compared with unvaccinated individuals/ reference period in general population.

Supplementary Figure S3. Contour-enhanced funnel plot for Dose 2 of mRNA-1273 compared with unvaccinated individuals/ reference period in general population.

Supplementary Table S1: PICOs investigated by included studies

Supplementary Table S2: Summary of meta-analysis results

Supplementary Table S3: Meta-analysis results by age subgroup

Supplementary Table S4: Meta-analysis results by risk window length subgroup

Supplementary Table S5: Meta-analysis results by study design subgroups

Supplementary Table S6: Meta-analysis results by overall risk of bias (RoB) subgroups

Supplementary Table S7: Meta-analysis results by confounding RoB subgroups

Supplementary Table S8: Meta-analysis results by outcome definition subgroups

Supplementary Table S9: Meta-analysis results by subgroups of approaches to handle COVID-19 infection in the analysis

## Data availability

The data used for the analysis and the analysis codes are available at: https://github.com/trangtph/Myocarditis_Vaccine_SR_public

## Author contributions

TT: Conceptualization, methodology, investigation, formal analysis, writing – original draft, visualisation, project administration; MB: conceptualisation, investigation, writing – review and editing, supervision; OK: conceptualisation, validation, writing – review and editing, supervision; SB: conceptualisation, investigation, writing – review and editing, supervision.

