## Supplementary Figures and Tables for "Quality and methodological heterogeneity of COVID-19 vaccine safety studies focusing on the myocarditis safety signal: A systematic review, meta-analysis and meta-regression"

**Figures:**

Supplementary Figure S1: PRISMA flow diagram

Supplementary Figure S2. Contour-enhanced funnel plot for Dose 2 of BNT-162b2 compared with unvaccinated individuals/ reference period in general population.

Supplementary Figure S3. Contour-enhanced funnel plot for Dose 2 of mRNA-1273 compared with unvaccinated individuals/ reference period in general population.

**Tables:**

Supplementary Table S1: PICOs investigated by included studies

Supplementary Table S2: Summary of meta-analysis results

Supplementary Table S3: Meta-analysis results by age subgroup

Supplementary Table S4: Meta-analysis results by risk window length subgroup

Supplementary Table S5: Meta-analysis results by study design subgroups

Supplementary Table S6: Meta-analysis results by overall risk of bias (RoB) subgroups

Supplementary Table S7: Meta-analysis results by confounding RoB subgroups

Supplementary Table S8: Meta-analysis results by outcome definition subgroups

Supplementary Table S9: Meta-analysis results by subgroups of approaches to handle COVID-19 infection in the analysis

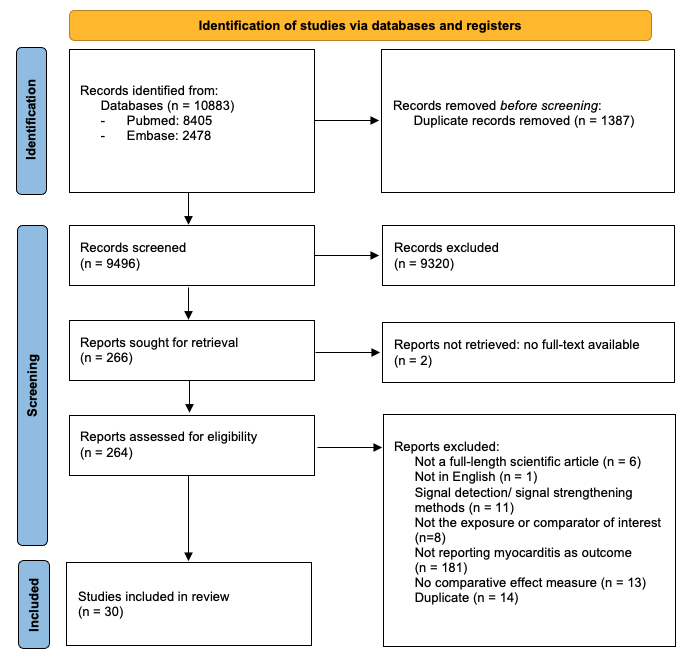

Figure S1. PRISMA flow diagram of study identification and inclusion

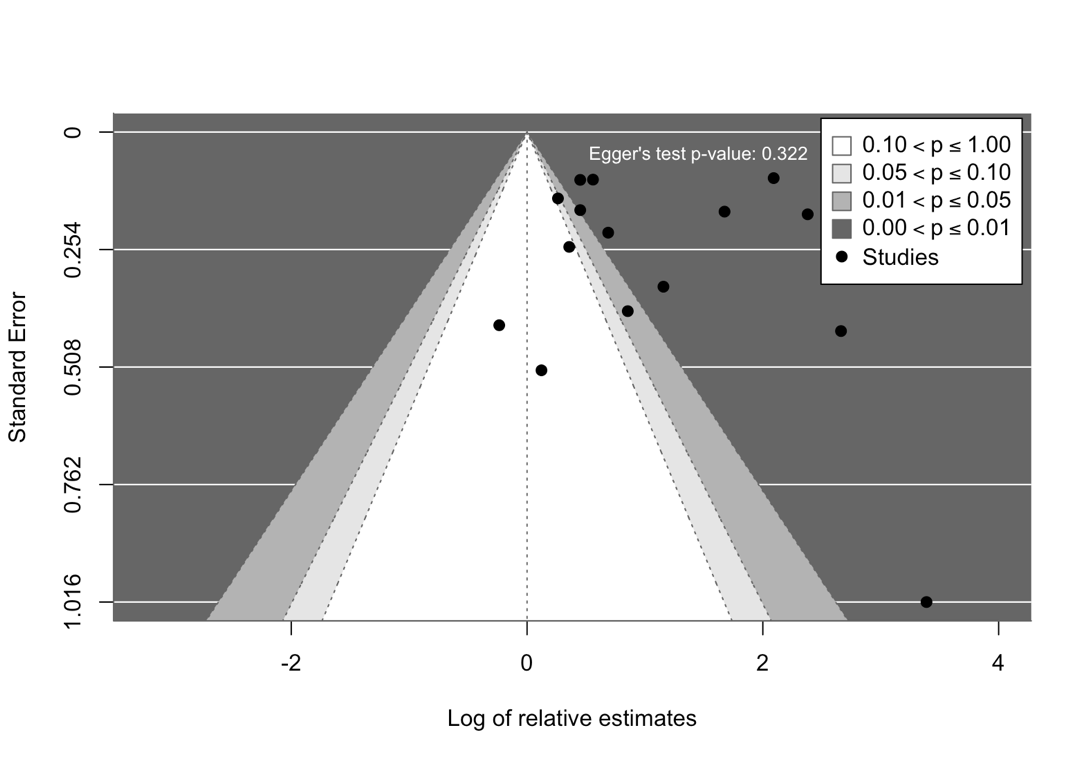

Figure S2. Contour-enhanced funnel plot for Dose 2 of BNT-162b2 compared with unvaccinated individuals/ reference period in general population. The Egger’s test for asymmetry and the distribution of studies in both non-significant (white) and significant (shaded) areas suggest that there is no strong evidence that publication bias is driving the results.

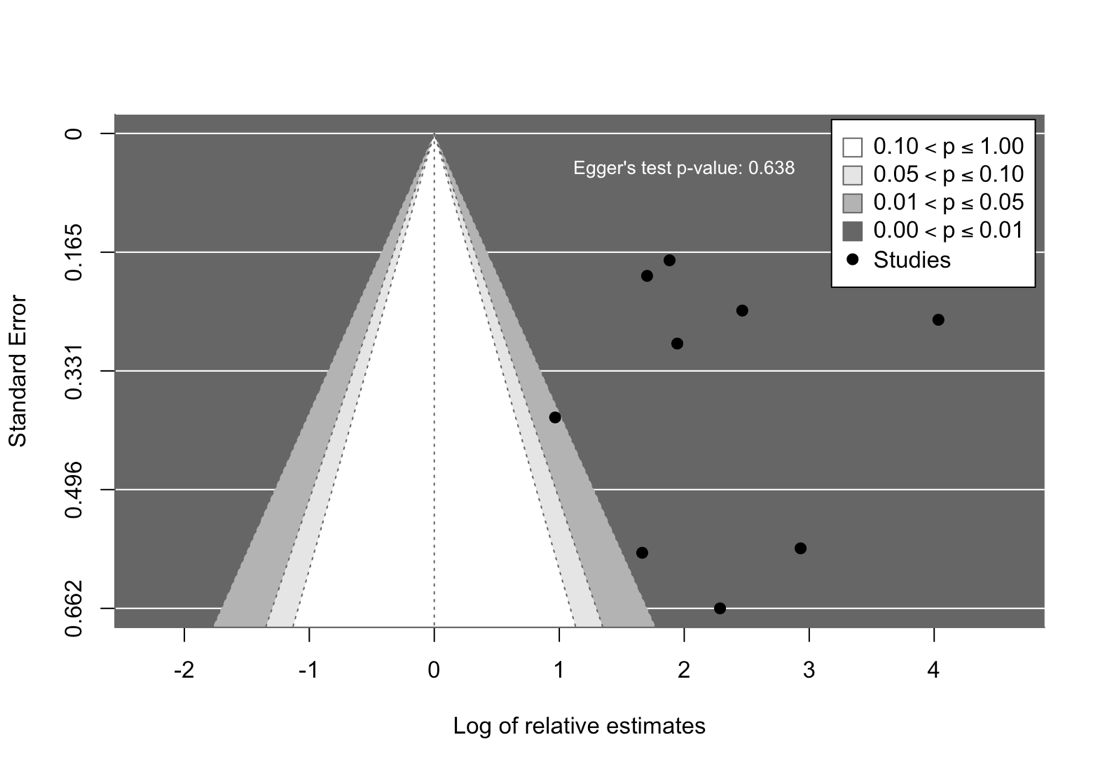

Figure S3. Contour-enhanced funnel plot for Dose 2 of mRNA-1273 compared with unvaccinated individuals/ reference period in general population. Most studies lied in the significant (shaded) areas, however there is no statistical evidence for funnel plot asymmetry (Egger’s test p-value > 0.05)

Table S1: PICOs investigated by 30 included studies

| Intervention | | Comparator | Population | Number of studies | Study |
| --- | --- | --- | --- | --- | --- |
| Vaccine | Dose |  |  |  |  |
| Ad26COVS1 | First dose | Unvaccinated individual/ reference time | General population | 1 | ^1^ |
| BNT-162b2 | First dose | Unvaccinated individual/ reference time | General population | 14 | ^1-14^ |
| BNT-162b2 | Second dose | Unvaccinated individual/ reference time | General population | 14 | ^1-12,14,15^ |
| BNT-162b2 | First and second doses | First and second dose of mRNA-1273 vaccine | Veteran | 1 | ^16^ |
| BNT-162b2 | First and second doses | Unvaccinated individual/ reference time | General population | 1 | ^17^ |
| BNT-162b2 | First or second dose | Unvaccinated individual/ reference time | General population | 3 | ^4,5,12^ |
| BNT-162b2 | Third dose | Unvaccinated individual/ reference time | General population | 5 | ^2,3,11,14,18^ |
| BNT-162b2 | Third dose | Unvaccinated individual/ reference time | High-risk of severe COVID-19 outcomes | 1 | ^19^ |
| BNT-162b2 | Fourth dose | Unvaccinated individual/ reference time | General population | 1 | ^20^ |
| BNT-162b2 | Fourth dose | Unvaccinated individual/ reference time | High-risk of severe COVID-19 outcomes | 1 | ^19^ |
| BNT-162b2 | Fourth dose | Unvaccinated individual/ reference time | Individuals aged >18 with particular comorbidities | 1 | ^21^ |
| BNT-162b2 | Third or fourth dose | Unvaccinated individual/ reference time | High-risk of severe COVID-19 outcomes | 1 | ^19^ |
| BNT-162b2 | Any dose | Unvaccinated individual/ reference time | General population | 1 | ^2^ |
| ChAdOx1 | First dose | Unvaccinated individual/ reference time | General population | 5 | ^1,2,10,11,14^ |
| ChAdOx1 | Second dose | Unvaccinated individual/ reference time | General population | 5 | ^1,2,10,11,14^ |
| ChAdOx1 | Any dose | Unvaccinated individual/ reference time | General population | 1 | ^2^ |
| CoronaVac | First dose | Unvaccinated individual/ reference time | General population | 3 | ^3,12,22^ |
| CoronaVac | First or second dose | Unvaccinated individual/ reference time | General population | 2 | ^12,23^ |
| CoronaVac | Second dose | Unvaccinated individual/ reference time | General population | 3 | ^3,12,22^ |
| CoronaVac | Third dose | Unvaccinated individual/ reference time | General population | 1 | ^3^ |
| mRNA-1273 | First dose | Unvaccinated individual/ reference time | General population | 10 | ^1,2,4-6,8-11,14^ |
| mRNA-1273 | First dose | First dose of BNT162b2 | General population | 1 | ^24^ |
| mRNA-1273 | Second dose | Unvaccinated individual/ reference time | General population | 10 | ^1,2,4-6,8-11,14^ |
| mRNA-1273 | Second dose | Second dose of BNT162b2 | General population | 3 | ^4,25,26^ |
| mRNA-1273 | Second dose | First or first or second doses of BNT162b2 | General population | 1 | ^4^ |
| mRNA-1273 | First or second dose | Unvaccinated individual/ reference time | General population | 2 | ^4,5^ |
| mRNA-1273 | First or second dose | First or first or second doses of BNT162b2 | General population | 1 | ^4^ |
| mRNA-1273 | Third dose | Unvaccinated individual/ reference time | General population | 3 | ^11,14,18^ |
| mRNA-1273 | Fourth dose | Unvaccinated individual/ reference time | General population | 1 | ^20^ |
| mRNA-1273 | Any dose | Unvaccinated individual/ reference time | General population | 1 | ^2^ |
| Bivalent mRNA boosters | Fourth dose | Unvaccinated individual/ reference time | General population | 1 | ^20^ |
| Heterologious booster: ChAdOx1-S - mRNA - mRNA | Third dose | Homologous booster: mRNA - mRNA - mRNA | General population | 1 | ^27^ |
| Heterologious primary: ChAdOx1-S - mRNA | Second dose | Homologous primary: mRNA - mRNA | General population | 1 | ^27^ |
| monovalent mRNA | Any dose | Unvaccinated individual/ reference time | General population | 1 | ^28^ |
| mRNA vaccines (BNT-162b2 or mRNA-1273) | First and second doses | Unvaccinated individual/ reference time | 1 (prevalent heart failure) | 1 | ^29^ |
| BNT-162b2, mRNA-1273 or Ad26COVS1 | Any dose | Unvaccinated individual/ reference time | General population | 1 | ^30^ |

Table S2: Summary of meta-analysis results of all PICOs, sorted by magnitude of I^2^

| PICO  (all PICOs were in general population) | Number of studies | Pooled effect estimate  [95% CI] | Prediction interval | Tau squared | I^2^ |
| --- | --- | --- | --- | --- | --- |
| BNT-162b2 Dose 2 Compared to Unvaccinated individual/time | 15 | 2.856  [1.665; 4.901] | [0.404; 20.198] | 0.762 | 95.6%  [94.1; 96.8] |
| mRNA-1273 Dose 2 Compared to Unvaccinated individual/time | 10 | 10.452  [5.376; 20.321] | [1.233; 88.63] | 0.769 | 91.7% [86.8; 94.8] |
| mRNA-1273 Dose 3 Compared to Unvaccinated individual/time | 4 | 2.179  [0.455; 10.432] | [0.025; 188.853] | 0.832 | 90.9% [79.9; 95.9] |
| BNT-162b2 Dose 1 or 2 Compared to Unvaccinated individual/time | 3 | 2.366  [0.246; 22.756] | [0; 779515.39] | 0.726 | 88.2% [67.2; 95.8] |
| mRNA-1273 Dose 2 Compared to Second dose of BNT162b2 | 3 | 2.658  [0.729; 9.69] | [0.002; 3421.797] | 0.226 | 83.8% [51.2; 94.6] |
| BNT-162b2 Dose 3 Compared to Unvaccinated individual/time | 6 | 1.899  [1.008; 3.578] | [0.399; 9.045] | 0.257 | 82.8% [63.6; 91.8] |
| ChAdOx1 Dose 1 Compared to Unvaccinated individual/time | 6 | 1.447  [0.661; 3.167] | [0.274; 7.635] | 0.29 | 69.9% [29.5; 87.1] |
| BNT-162b2 Dose 1 Compared to Unvaccinated individual/time | 15 | 1.726  [1.375; 2.166] | [0.844; 3.53] | 0.098 | 65.4% [40.2; 80] |
| mRNA-1273 Dose 1 Compared to Unvaccinated individual/time | 11 | 2.759  [1.801; 4.225] | [0.835; 9.11] | 0.238 | 60.4% [23.2; 79.5] |
| CoronaVac Dose 1 Compared to Unvaccinated individual/time | 3 | 1.337  [0.206; 8.688] | [0.001; 1986.974] | 0.156 | 32.7%  [0; 93] |
| ChAdOx1 Dose 2 Compared to Unvaccinated individual/time | 6 | 0.959  [0.756; 1.217] | [0.767; 1.2] | 0 | 24.4%  [0; 68.1] |
| CoronaVac Dose 2 Compared to Unvaccinated individual/time | 3 | 1.012  [0.241; 4.253] | [0.005; 194.775] | 0 | 0%  [0; 89.6] |

Table S3: Meta-analysis results by age subgroup. General population: when the study did not have age restriction; Older population: when the study was restricted to individuals aged >50 years; Younger population: when the study was restricted to individuals aged <40 years.

| **PICO** | **N study** | **Pooled effect estimate [95% CI]** | | | **p-value subgroup difference** | **N study in each subgroup (general – older – younger)** |
| --- | --- | --- | --- | --- | --- | --- |
|  |  | **General population** | **Older population** | **Younger population** |  |  |
| BNT-162b2 Dose 1  Comparator: Unvaccinated individual/time | 15 | 1.697  [1.3; 2.214] |  | 1.938  [0.892; 4.207] | 0.662 | 10 - 0 - 5 |
| BNT-162b2 Dose 1 or 2  Comparator: Unvaccinated individual/time | 3 | 1.38  [0.719; 2.649] |  | 6.94  [3.488; 13.807] | <0.001 | 2 - 0 - 1 |
| BNT-162b2 Dose 2  Comparator: Unvaccinated individual/time | 15 | 2.226  [1.283; 3.863] |  | 6.657  [1.135; 39.034] | 0.072 | 11 - 0 - 4 |
| BNT-162b2 Dose 3  Comparator: Unvaccinated individual/time | 6 | 1.692  [1.159; 2.47] | 1.13  [0.931; 1.371] | 4.38  [2.595; 7.393] | <0.001 | 4 - 1 - 1 |
| ChAdOx1 Dose 1  Comparator: Unvaccinated individual/time | 6 | 1.295  [1.079; 1.553] |  | 5.23  [2.482; 11.02] | <0.001 | 5 - 0 - 1 |
| ChAdOx1 Dose 2  Comparator: Unvaccinated individual/time | 6 | 0.958  [0.718; 1.28] |  | 1.01  [0.249; 4.105] | 0.942 | 5 - 0 - 1 |
| CoronaVac Dose 1  Comparator: Unvaccinated individual/time | 3 | 1.018  [0; 6142.747] | 1.96  [0.697; 5.509] |  | 0.448 | 2 - 1 - 0 |
| CoronaVac Dose 2  Comparator: Unvaccinated individual/time | 3 | 0.682  [0.57; 0.818] | 1.78  [0.502; 6.308] |  | 0.138 | 2 - 1 - 0 |
| mRNA-1273 Dose 1  Comparator: Unvaccinated individual/time | 11 | 2.331  [1.461; 3.718] |  | 4.135  [0.682; 25.084] | 0.216 | 8 - 0 - 3 |
| mRNA-1273 Dose 2  Comparator: Second dose of BNT162b2 | 3 | 2.78  [1.671; 4.624] |  | 2.54  [0.004; 1735.534] | 0.875 | 1 - 0 - 2 |
| mRNA-1273 Dose 2  Comparator: Unvaccinated individual/time | 10 | 9.267  [5.118; 16.778] |  | 14.223  [0.287; 705.684] | 0.648 | 7 - 0 - 3 |
| mRNA-1273 Dose 3  Comparator: Unvaccinated individual/time | 4 | 1.669  [0.002; 1629.323] | 1.13  [0.926; 1.379] | 7.88  [4.021; 15.443] | <0.001 | 2 - 1 - 1 |

Table S4: Meta-analysis results by risk window length subgroups

| **PICO** | **N study** | **Pooled effect estimate [95% CI]** | | | **p-value subgroup difference** | **N study in each subgroup (7/14 – 21/28 – others)** |
| --- | --- | --- | --- | --- | --- | --- |
|  |  | **Risk window 7-day or 14-day post vaccination** | **Risk window 21-day or 28-day post vaccination** | **Other risk window lengths** |  |  |
| BNT-162b2 Dose 1  Comparator: Unvaccinated individual/time | 15 | 2.563  [1.552; 4.234] | 1.413  [1.264; 1.579] | 1.61  [0.292; 8.889] | 0.006 | 5 - 9 - 1 |
| BNT-162b2 Dose 1 or 2  Comparator: Unvaccinated individual/time | 3 | 6.94  [3.488; 13.807] | 1.38  [0.719; 2.649] |  | <0.001 | 1 - 2 - 0 |
| BNT-162b2 Dose 2  Comparator: Unvaccinated individual/time | 15 | 5.691  [1.428; 22.684] | 1.644  [1.38; 1.959] |  | 0.014 | 5 - 10 - 0 |
| BNT-162b2 Dose 3  Comparator: Unvaccinated individual/time | 6 | 2.9  [0.562; 14.953] | 1.355  [0.714; 2.569] |  | 0.063 | 3 - 3 - 0 |
| ChAdOx1 Dose 1  Comparator: Unvaccinated individual/time | 6 | 2.707  [0.001; 9350.38] | 1.287  [1.01; 1.641] |  | 0.25 | 2 - 4 - 0 |
| ChAdOx1 Dose 2  Comparator: Unvaccinated individual/time | 6 | 0.612  [0.02; 18.457] | 0.986  [0.724; 1.342] |  | 0.094 | 2 - 4 - 0 |
| CoronaVac Dose 1  Comparator: Unvaccinated individual/time | 3 | 0.5  [0.131; 1.906] | 1.964  [1.903; 2.028] |  | 0.045 | 1 - 2 - 0 |
| CoronaVac Dose 2  Comparator: Unvaccinated individual/time | 3 | 0.69  [0.182; 2.622] | 1.267  [0.003; 468.992] |  | 0.461 | 1 - 2 - 0 |
| mRNA-1273 Dose 1  Comparator: Unvaccinated individual/time | 11 | 4.887  [2.022; 11.813] | 2.078  [1.334; 3.237] |  | 0.01 | 4 - 7 - 0 |
| mRNA-1273 Dose 2  Comparator: Second dose of BNT162b2 | 3 | 2.54  [0.004; 1735.534] | 2.78  [1.671; 4.624] |  | 0.875 | 2 - 1 - 0 |
| mRNA-1273 Dose 2  Comparator: Unvaccinated individual/time | 10 | 35.307  [10.382; 120.069] | 6.448  [4.246; 9.792] |  | <0.001 | 3 - 7 - 0 |
| mRNA-1273 Dose 3  Comparator: Unvaccinated individual/time | 4 | 2.773  [0; 3052642.445] | 1.595  [0.008; 317.881] |  | 0.637 | 2 - 2 - 0 |

Table S5: Meta-analysis results by study design subgroups

| **PICO** | **N study** | **Pooled effect estimate [95% CI]** | | | **p-value subgroup difference** | **N study in each subgroup (Cohort – SCCS/SCRI - Others)** |
| --- | --- | --- | --- | --- | --- | --- |
|  |  | **Cohort** | **SCCS/SCRI** | **Other designs** |  |  |
| BNT-162b2 Dose 1  Comparator: Unvaccinated individual/time | 15 | 1.422  [1.061; 1.904] | 1.785  [1.267; 2.515] | 1.885  [0.289; 12.274] | 0.181 | 4 - 9 - 2 |
| BNT-162b2 Dose 1 or 2  Comparator: Unvaccinated individual/time | 3 | 1.34  [0.899; 1.998] | 1.51  [0.749; 3.045] | 6.94  [3.488; 13.807] | <0.001 | 1 - 1 - 1 |
| BNT-162b2 Dose 2  Comparator: Unvaccinated individual/time | 15 | 1.723  [1.223; 2.428] | 2.343  [1.127; 4.868] | 9.244  [0.433; 197.286] | <0.001 | 5 - 8 - 2 |
| BNT-162b2 Dose 3  Comparator: Unvaccinated individual/time | 6 | 1.1  [0.341; 3.549] | 2.031  [0.944; 4.37] |  | 0.352 | 1 - 5 - 0 |
| ChAdOx1 Dose 1  Comparator: Unvaccinated individual/time | 6 | 0.25  [0.037; 1.686] | 1.595  [0.756; 3.362] |  | 0.067 | 1 - 5 - 0 |
| ChAdOx1 Dose 2  Comparator: Unvaccinated individual/time | 6 | 0.76  [0.24; 2.408] | 0.963  [0.723; 1.284] |  | 0.691 | 1 - 5 - 0 |
| mRNA-1273 Dose 1  Comparator: Unvaccinated individual/time | 11 | 1.78  [0.344; 9.219] | 3.492  [1.755; 6.951] | 3.13  [1.367; 7.163] | 0.313 | 3 - 6 - 2 |
| mRNA-1273 Dose 2  Comparator: Second dose of BNT162b2 | 3 | 3.61  [0.326; 39.951] |  | 1.48  [0.878; 2.495] | 0.006 | 2 - 0 - 1 |
| mRNA-1273 Dose 2  Comparator: Unvaccinated individual/time | 10 | 6.215  [4.626; 8.35] | 10.221  [2.376; 43.973] | 28.746  [5.166; 159.966] | <0.001 | 3 - 5 - 2 |

Table S6: Meta-analysis results by overall risk of bias (RoB) subgroups

| **PICO** | **N study** | **Pooled effect estimate [95% CI]** | | **p-value subgroup difference** | **N study in each subgroup (Low/moderate – Serious/critical)** |
| --- | --- | --- | --- | --- | --- |
|  |  | **Low – moderate RoB** | **Serious – critical RoB** |  |  |
| BNT-162b2 Dose 1  Comparator: Unvaccinated individual/time | 15 | 1.673  [1.27; 2.203] | 1.763  [1.139; 2.729] | 0.777 | 11 - 4 |
| BNT-162b2 Dose 1 or 2  Comparator: Unvaccinated individual/time | 3 | 6.94  [3.488; 13.807] | 1.38  [0.719; 2.649] | <0.001 | 2 - 1 |
| BNT-162b2 Dose 2  Comparator: Unvaccinated individual/time | 15 | 5.629  [1.329; 23.836] | 2.171  [1.237; 3.81] | 0.098 | 10 - 5 |
| BNT-162b2 Dose 3  Comparator: Unvaccinated individual/time | 6 | 1.1  [0.341; 3.549] | 2.031  [0.944; 4.37] | 0.352 | 5 - 1 |
| ChAdOx1 Dose 1  Comparator: Unvaccinated individual/time | 6 | 1.595  [0.756; 3.362] | 0.25  [0.037; 1.686] | 0.067 | 5 - 1 |
| ChAdOx1 Dose 2  Comparator: Unvaccinated individual/time | 6 | 0.963  [0.723; 1.284] | 0.76  [0.24; 2.408] | 0.691 | 5 - 1 |
| mRNA-1273 Dose 1  Comparator: Unvaccinated individual/time | 11 | 3.358  [2.677; 4.211] | 2.594  [1.375; 4.895] | 0.346 | 8 - 3 |
| mRNA-1273 Dose 2  Comparator: Second dose of BNT162b2 | 3 | 2.54  [0.004; 1735.534] | 2.78  [1.671; 4.624] | 0.875 | 1 - 2 |
| mRNA-1273 Dose 2  Comparator: Unvaccinated individual/time | 10 | 14.243  [1.477; 137.354] | 9.088  [3.694; 22.355] | 0.484 | 7 - 3 |

Table S7: Meta-analysis results by confounding RoB subgroups

| **PICO** | **N study** | **Pooled effect estimate [95% CI]** | | **p-value subgroup difference** | **N study in each subgroup (Low/moderate – Serious/critical)** |
| --- | --- | --- | --- | --- | --- |
|  |  | **Low – moderate RoB** | **Serious – critical RoB** |  |  |
| BNT-162b2 Dose 1  Comparator: Unvaccinated individual/time | 15 | 1.673  [1.27; 2.203] | 1.763  [1.139; 2.729] | 0.777 | 11 - 4 |
| BNT-162b2 Dose 1 or 2 Comparator: Unvaccinated individual/time | 3 | 6.94  [3.488; 13.807] | 1.38  [0.719; 2.649] | <0.0001 | 2 - 1 |
| BNT-162b2 Dose 2  Comparator: Unvaccinated individual/time | 15 | 5.629  [1.329; 23.836] | 2.171  [1.237; 3.81] | 0.098 | 10 - 5 |
| BNT-162b2 Dose 3  Comparator: Unvaccinated individual/time | 6 | 1.1  [0.341; 3.549] | 2.031  [0.944; 4.37] | 0.352 | 5 - 1 |
| ChAdOx1 Dose 1  Comparator: Unvaccinated individual/time | 6 | 1.595  [0.756; 3.362] | 0.25  [0.037; 1.686] | 0.067 | 5 - 1 |
| ChAdOx1 Dose 2  Comparator: Unvaccinated individual/time | 6 | 0.963  [0.723; 1.284] | 0.76  [0.24; 2.408] | 0.691 | 5 - 1 |
| mRNA-1273 Dose 1  Comparator: Unvaccinated individual/time | 11 | 3.358  [2.677; 4.211] | 2.594  [1.375; 4.895] | 0.346 | 8 - 3 |
| mRNA-1273 Dose 2  Comparator: Second dose of BNT162b2 | 3 | 2.54  [0.004; 1735.534] | 2.78  [1.671; 4.624] | 0.875 | 1 - 2 |
| mRNA-1273 Dose 2  Comparator: Unvaccinated individual/time | 10 | 14.243  [1.477; 137.354] | 9.088  [3.694; 22.355] | 0.484 | 7 - 3 |

Table S8: Meta-analysis results by outcome definition subgroups

| **PICO** | **N study** | **Pooled effect estimate [95% CI]** | | **p-value subgroup difference** | **N study in each subgroup (Myo – Myo/peri)** |
| --- | --- | --- | --- | --- | --- |
|  |  | **Myocarditis only** | **Myocarditis or pericarditis** |  |  |
| BNT-162b2 Dose 1 Comparator: Unvaccinated individual/time Population 0 | 15 | 1.478  [1.29; 1.693] | 1.962  [1.276; 3.018] | 0.136 | 7 - 8 |
| BNT-162b2 Dose 2 Comparator: Unvaccinated individual/time Population 0 | 15 | 2.591  [1.264; 5.311] | 3.051  [1.071; 8.694] | 0.756 | 8 - 7 |
| BNT-162b2 Dose 3 Comparator: Unvaccinated individual/time Population 0 | 6 | 1.685  [0.515; 5.514] | 2.202  [0.696; 6.965] | 0.474 | 2 - 4 |
| ChAdOx1 Dose 1 Comparator: Unvaccinated individual/time Population 0 | 6 | 2.707  [0.001; 9350.38] | 1.287  [1.01; 1.641] | 0.25 | 4 - 2 |
| ChAdOx1 Dose 2 Comparator: Unvaccinated individual/time Population 0 | 6 | 0.986  [0.724; 1.342] | 0.612  [0.02; 18.457] | 0.094 | 4 - 2 |
| CoronaVac Dose 1 Comparator: Unvaccinated individual/time Population 0 | 3 | 1.018  [0; 6142.747] | 1.96  [0.697; 5.509] | 0.448 | 1 - 2 |
| CoronaVac Dose 2 Comparator: Unvaccinated individual/time Population 0 | 3 | 0.682  [0.57; 0.818] | 1.78  [0.502; 6.308] | 0.138 | 1 - 2 |
| mRNA-1273 Dose 1 Comparator: Unvaccinated individual/time Population 0 | 11 | 2.316  [1.4; 3.831] | 3.531  [1.271; 9.811] | 0.311 | 6 - 5 |
| mRNA-1273 Dose 2 Comparator: Second dose of BNT162b2 Population 0 | 3 | 4.14  [3.184; 5.383] | 2.034  [0.037; 111.605] | 0.038 | 2 - 1 |
| mRNA-1273 Dose 2 Comparator: Unvaccinated individual/time Population 0 | 10 | 11.867  [1.429; 98.529] | 9.703  [4.708; 19.996] | 0.78 | 6 - 4 |
| mRNA-1273 Dose 3 Comparator: Unvaccinated individual/time Population 0 | 4 | 2.031  [0.104; 39.755] | 2.64  [1.25; 5.578] | 0.74 | 1 - 3 |

Table S9: Meta-analysis results by subgroups of approaches to handle COVID-19 infection in the analysis. Approaches that measure direct effect of vaccination include censoring COVID-19 cases, excluding COVID-19 cases before and during the study. Other approaches include excluding COVID-19 cases and COVID-19 as covariate in the model.

| **PICO** | **N study** | **Pooled effect estimate [95% CI]** | | | **p-value subgroup difference** | **N study in each subgroup (No approach – Direct effect – Others)** |
| --- | --- | --- | --- | --- | --- | --- |
|  |  | **No approach applied** | **Approaches that measure direct effect of vaccination only** | **Others approaches** |  |  |
| BNT-162b2 Dose 1  Comparator: Unvaccinated individual/time | 15 | 1.484  [1.283; 1.717] | 1.999  [0.355; 11.242] | 1.885  [0.289; 12.274] | 0.27 | 10 - 3 - 2 |
| BNT-162b2 Dose 1 or 2  Comparator: Unvaccinated individual/time | 3 | 1.51  [0.749; 3.045] | 1.34  [0.899; 1.998] | 6.94  [3.488; 13.807] | <0.0001 | 1 - 1 - 1 |
| BNT-162b2 Dose 2  Comparator: Unvaccinated individual/time | 15 | 2.055  [1.204; 3.508] | 3.01  [0.189; 47.856] | 9.244  [0.433; 197.286] | <0.0001 | 10 - 3 - 2 |
| BNT-162b2 Dose 3  Comparator: Unvaccinated individual/time | 6 | 1.991  [0.809; 4.9] | 4.72  [1.398; 15.941] | 1.13  [0.931; 1.371] | 0.016 | 4 - 1 - 1 |
| CoronaVac Dose 1  Comparator: Unvaccinated individual/time | 3 | 1.964  [1.903; 2.028] | 0.5  [0.131; 1.906] |  | 0.045 | 2 - 1 - 0 |
| CoronaVac Dose 2  Comparator: Unvaccinated individual/time | 3 | 1.267  [0.003; 468.992] | 0.69  [0.182; 2.622] |  | 0.461 | 2 - 1 - 0 |
| mRNA-1273 Dose 1  Comparator: Unvaccinated individual/time | 11 | 3.465  [2.037; 5.892] | 1.162  [1.116; 1.209] | 3.13  [1.367; 7.163] | <0.0001 | 7 - 2 - 2 |
| mRNA-1273 Dose 2  Comparator: Second dose of BNT162b2 | 3 | 3.61  [0.326; 39.951] |  | 1.48  [0.878; 2.495] | 0.006 | 2 - 0 - 1 |
| mRNA-1273 Dose 2  Comparator: Unvaccinated individual/time | 10 | 9.159  [2.958; 28.359] | 6.68  [4.715; 9.463] | 28.746  [5.166; 159.966] | <0.0001 | 6 - 2 - 2 |
| mRNA-1273 Dose 3  Comparator: Unvaccinated individual/time | 4 | 2.823  [0.193; 41.341] |  | 1.13  [0.926; 1.379] | 0.147 | 3 - 0 - 1 |
