## Supplementary file 1 for "Quality and methodological heterogeneity of COVID-19 vaccine safety studies focusing on the myocarditis safety signal: A systematic review, meta-analysis and meta-regression"

### Methodology of COVID-19 Vaccine Safety study – Systematic Review protocol

Version history

| Version date | Changes made |
|---|---|
| October 10 2023 | First version of systematic review protocol |
| April 20 2024 | Data extraction: extract outcome data for all papers and full data extractions for 3 most common outcomes |
| June 14 2024 | First version of meta-analysis and meta-regression protocol |
| July 18, 2024 | Data extraction: full data extraction for only myocarditis and Bell's palsy outcomes<br>Meta-analysis: add decision rule for selection of effect estimates |
| August 12, 2024 | Data analysis: change from multi-level model to robust variance estimation approach |
| October 3, 2024 | Add post-hoc analysis |

#### 1 Background

The rapid development and distribution of COVID-19 vaccines marked a major scientific achievement. However, their accelerated approval underscored the need for postmarketing safety monitoring to detect rare adverse events, such as myocarditis. While multiple real-world studies confirmed an increased risk of myocarditis after vaccination, reported effect estimates varied widely. This heterogeneity complicates benefit-risk assessments and public confidence in vaccine safety.

Variability in study findings often arises from clinical and methodological differences. Clinical diversity includes differences in participant characteristics, vaccination policies, and healthcare access, affecting how results apply to different populations. Methodological differences stem from variations in study design, follow-up duration, comparator selection, and bias control. Designing vaccine safety studies faces unique challenges, such as seasonal vaccination patterns and high coverage, making it difficult to identify appropriate control groups. Specialized methods like case-centred and self-controlled designs address these issues but rely on strict assumptions.

Prior research suggests study design choices contribute to effect estimate variability, though findings may not fully apply to the COVID-19 pandemic, given healthcare constraints and strong time trends in vaccination. To better understand these challenges, we will examine the methodologies of comparative observational studies assessing myocarditis risk after COVID-19 vaccination.

### 2 Objectives

This project aims to provide an overview of methodologies of inferential, comparative observational studies that evaluate COVID-19 vaccine safety signals. The specific objectives include:

(1) To describe and assess the quality of the current practice in study design and analytical approaches used in observational studies on COVID-19 vaccine safety
(2) To assess the heterogeneity in findings across studies that addressed similar safety questions and investigate the impact of study design and analysis choice on the observed heterogeneity of those studies

### 3 Methods

#### 3.1 Study design

A methodological systematic review of inferential, comparative observational studies investigating safety of COVID-19 vaccines will be conducted. Statistical heterogeneity will be quantified, and subsequent meta-analyses will be carried out where applicable. Possible approaches such as meta-regression will be employed to investigate sources of heterogeneity. This systematic review will be performed conforming to the Preferred Reporting Items for Systematic Reviews and Meta-Analyses 2020 (PRISMA) (1).

#### 3.2 Eligibility criteria

Studies will be included in our systematic review if all following criteria are fulfilled:

1) Observational study or study protocol that aims to quantify the safety of COVID-19 vaccines among individuals vaccinated with COVID-19 vaccines.
2) Include an exposed group of individuals (in parallel-group design) or time period (in self-controlled design) that receive (at least one dose of) COVID-19 vaccines and a control group of individuals or time period that are not exposed to COVID-19 vaccines, or have another COVID-19 vaccination patterns.
3) Compare the effect of COVID-19 vaccines on a safety outcome between an exposed group and a control group/ control period and report the comparative effect measurement (ratio or difference) between groups.
4) Published in English as a peer-reviewed scientific article.
5) Published after December 11th, 2020 (first COVID-19 vaccine emergency use authorization) (2)
6) Full-text available

Studies with primary objectives of safety signal detection (which employ data mining technique without prior hypothesis), and signal strengthening (such as disproportionality analysis, ecological, and crude before-after comparison) will be excluded, due to difference in rigor requirement of study conduct and report. Editorials, letters to editor and commentaries will also be excluded.

#### 3.3 Search strategy

Structural searches will be conducted on Pubmed, Embase and Web of Science to identify relevant records. The search strategies will combine keywords and subject heading terms related to (1) COVID-19 vaccines, (2) safety, (3) comparison of effect measurement, using filters to restrict search to English articles, studies on human subjects, studies that are not clinical trial, case report, and review, and studies that are published after December 11, 2020. The search string is structured as follow: (((Block 1A AND 1B, search within 5 words proximity) OR (1C AND 1B) OR 1D) AND 2 AND 3) NOT 4. The European Union electronic Register of Post-Authorisation Studies (EU PAS Register) and the database ClinicalTrials.gov will also be searched, using keywords "COVID-19", "vaccine", filter "observational study". The reference lists of eligible studies will be screened for additional eligible papers.

*Table 1. Search strategy*

| **1) COVID-19 vaccines** | | **2) Safety** | **3) Comparison of effect measurement** | **4) Study design exclusion** |
|---|---|---|---|---|
| **1A) COVID-19:** | **1B) Vaccine:** | Safety | Associate* | "Case Reports" [Publication Type] |
| | Vaccin* | Harm* | Association* | |
| SARS-CoV-2 | | Adverse event* | | "Clinical Trial, Phase I" [Publication Type] |
| COVID-19 | "vaccin*"[Title/Abstract] | Adverse effect* | Comparing | |
| nCoV-19 | | Adverse drug reaction* | Compare* | |
| 2019-nCoV | | Adverse drug event* | Comparative | "Clinical Trial, Phase II" [Publication Type] |
| Coronavirus | | Adverse reaction* | Comparison | |
| | | | Versus | |
| "SARS-CoV-2"[Mesh] | | Side effect* | | "Clinical Trial, Phase III" [Publication Type] |
| | | Complication* | Risk ratio* | |
| "COVID-19"[Mesh] | | Undesirable effect* | RR | "Randomized Controlled Trial" [Publication Type] |
| | | Unfavorable effect* | Risk difference* | |
| | | | Relative risk | |
| "SARS-CoV-2"[Title/Abstract] OR "COVID-19"[Title/Abstract] OR "nCoV-19"[Title/Abstract] OR "2019-nCoV"[Title/Abstract] OR "Coronavirus"[Title/Abstract] OR "SARS-CoV-2"[MeSH Terms] OR "COVID-19"[MeSH Terms] | | Pharmacovigilance | Absolute risk* | |
| | | | Relative incidence | |
| | | Risk* | | |
| | | | Odds ratio* | |
| | | Drug-Related Side Effects and Adverse Reactions [Mesh] | aOR | |
| | | | Incidence rate ratio* | |
| | | "COVID-19 Vaccines/adverse effects"[Mesh] | IRR | |
| | | | IRRs | |
| | | "Safety"[Title/Abstract] OR "harm*"[Title/Abstract] OR "adverse event*"[Title/Abstract] OR "adverse effect*"[Title/Abstract] OR "adverse drug reaction*"[Title/Abstract] OR "adverse drug event*"[Title/Abstract] OR "adverse reaction*"[Title/Abstract] OR "side effect*"[Title/Abstract] OR "complication*"[Title/Abstract] OR "undesirable effect*"[Title/Abstract] OR "unfavorable effect*"[Title/Abstract] OR "Pharmacovigilance"[Title/Abstract] OR "risk*"[Title/Abstract] OR "drug related side | Rate difference*<br>Hazard ratio*<br>HR<br>Relative rate<br>Attributable fraction | |

| | | | |
|---|---|---|---|
| | | effects and adverse reactions"[MeSH Terms] OR "covid 19 vaccines/adverse effects"[MeSH Terms] | Attributable risk<br>Attributable rate<br>Number needed to harm<br><br>NNH<br><br>Excess events<br>Excess cases<br>Excess number*<br><br>"Comparative Study"[Publication Type] |
| **1C) COVID-19 Vaccine brand:**<br>Pfizer-BioNTech<br>Comirnaty<br>BNT162b2<br>BNT162<br>BNT-162B2<br><br>mRNA-1273<br>Moderna<br>Spikevax<br><br>Ad26COVS1<br>Ad26.COV2.S<br>Jcovden<br>Janssen<br><br>ChAdOx1<br>ChAdOx1-S<br>Vaxzevria<br>AstraZeneca<br>Oxford/AstraZeneca<br>AZD1222<br><br>Sinovac<br>CoronaVac<br>Sinovac-CoronaVac<br><br>Novavax<br>Nuvaxovid<br><br>Bimervax | | | "associate*"[Title/Abstract] OR "association*"[Title/Abstract] OR "Comparing"[Title/Abstract] OR "compare*"[Title/Abstract] OR "Comparative"[Title/Abstract] OR "Comparison"[Title/Abstract] OR "Versus"[Title/Abstract] OR "risk ratio*"[Title/Abstract] OR "RR"[Title/Abstract] OR "risk difference*"[Title/Abstract] OR "relative risk"[Title/Abstract] OR "absolute risk*"[Title/Abstract] OR "relative incidence"[Title/Abstract] OR "odds ratio*"[Title/Abstract] OR "aOR"[Title/Abstract] OR "incidence rate ratio*"[Title/Abstract] OR "IRR"[Title/Abstract] OR "IRRs"[Title/Abstract] OR "rate difference*"[Title/Abstract] OR "hazard ratio*"[Title/Abstract] OR "HR"[Title/Abstract] OR "relative rate"[Title/Abstract] OR "attributable fraction"[Title/Abstract] OR "attributable risk"[Title/Abstract] OR "attributable rate"[Title/Abstract] OR "number needed to harm"[Title/Abstract] OR "NNH"[Title/Abstract] OR "excess events"[Title/Abstract] OR "excess cases"[Title/Abstract] OR "excess number*"[Title/Abstract] OR "comparative study"[Publication Type] |

| |
|---|
| HIPRA<br><br>PHH-1V<br><br><br>Sinopharm<br><br><br>Valneva<br><br>VLA2001<br><br><br>"Pfizer-BioNTech"[Title/Abstract] OR "Comirnaty"[Title/Abstract] OR "BNT162b2"[Title/Abstract] OR "BNT162"[Title/Abstract] OR "BNT-162B2"[Title/Abstract] OR "mRNA-1273"[Title/Abstract] OR "Moderna"[Title/Abstract] OR "Spikevax"[Title/Abstract] OR "Ad26COVS1"[Title/Abstract] OR "Ad26.COV2.S"[Title/Abstract] OR "Jcovden"[Title/Abstract] OR "Janssen"[Title/Abstract] OR "ChAdOx1"[Title/Abstract] OR "ChAdOx1-S"[Title/Abstract] OR "Vaxzevria"[Title/Abstract] OR "AstraZeneca"[Title/Abstract] OR "oxford astrazeneca"[Title/Abstract] OR "AZD1222"[Title/Abstract] OR "Sinovac"[Title/Abstract] OR "CoronaVac"[Title/Abstract] OR "Sinovac-CoronaVac"[Title/Abstract] OR "Novavax"[Title/Abstract] OR "Nuvaxovid"[Title/Abstract] OR "Bimervax"[Title/Abstract] OR "HIPRA"[Title/Abstract] OR "PHH-1V"[Title/Abstract] OR "Sinopharm"[Title/Abstract] OR "Valneva"[Title/Abstract] OR "VLA2001"[Title/Abstract] |
| **1D) COVID-19 Vaccine in general:**<br><br><br><br>COVID-19 Vaccines [MeSH Term]<br><br><br>"SARS-CoV-2 vaccine"[Title/Abstract:~5] OR "SARS-CoV-2 vaccines"[Title/Abstract:~5] OR "SARS-CoV-2 vaccination"[Title/Abstract:~5] OR "SARS-CoV-2 vaccinated"[Title/Abstract:~5] OR "COVID-19 vaccine"[Title/Abstract:~5] OR "COVID-19 vaccines"[Title/Abstract:~5] OR "COVID-19 vaccination"[Title/Abstract:~5] OR "COVID-19 vaccinated"[Title/Abstract:~5] OR "nCov-19 vaccine"[Title/Abstract:~5] OR "nCov-19 vaccines"[Title/Abstract:~5] OR "nCov-19 vaccination"[Title/Abstract:~5] OR "nCov-19 vaccinated"[Title/Abstract:~5] OR "2019-nCoV vaccine"[Title/Abstract:~5] OR "2019-nCoV vaccines"[Title/Abstract:~5] OR "2019-nCoV vaccination"[Title/Abstract:~5] OR "2019-nCoV vaccinated"[Title/Abstract:~5] OR "Coronavirus vaccine"[Title/Abstract:~5] OR "Coronavirus vaccines"[Title/Abstract:~5] OR "Coronavirus vaccination"[Title/Abstract:~5] OR "Coronavirus vaccinated"[Title/Abstract:~5] OR (("Pfizer-BioNTech"[Title/Abstract] OR "Comirnaty"[Title/Abstract] OR "BNT162b2"[Title/Abstract] OR "BNT162"[Title/Abstract] OR "BNT-162B2"[Title/Abstract] OR "mRNA-1273"[Title/Abstract] OR "Moderna"[Title/Abstract] OR "Spikevax"[Title/Abstract] OR "Ad26COVS1"[Title/Abstract] OR "Ad26.COV2.S"[Title/Abstract] OR "Jcovden"[Title/Abstract] OR "Janssen"[Title/Abstract] |

| | | | |
|---|---|---|---|
| OR "ChAdOx1"[Title/Abstract] OR "ChAdOx1-S"[Title/Abstract] OR "Vaxzevria"[Title/Abstract] OR "AstraZeneca"[Title/Abstract] OR "oxford astrazeneca"[Title/Abstract] OR "AZD1222"[Title/Abstract] OR "Sinovac"[Title/Abstract] OR "CoronaVac"[Title/Abstract] OR "Sinovac-CoronaVac"[Title/Abstract] OR "Novavax"[Title/Abstract] OR "Nuvaxovid"[Title/Abstract] OR "Bimervax"[Title/Abstract] OR "HIPRA"[Title/Abstract] OR "PHH-1V"[Title/Abstract] OR ("VidPrevtyn"[All Fields] AND "Beta"[Title/Abstract]) OR "Sinopharm"[Title/Abstract] OR "Valneva"[Title/Abstract] OR "VLA2001"[Title/Abstract]) AND "vaccin*"[Title/Abstract]) OR "COVID-19 vaccines"[MeSH Terms] | | | |
| Final search strings: | | | |
| **Pubmed:**<br><br>((("SARS-CoV-2 vaccine"[Title/Abstract:~3] OR "SARS-CoV-2 vaccines"[Title/Abstract:~3] OR "SARS-CoV-2 vaccination"[Title/Abstract:~3] OR "SARS-CoV-2 vaccinated"[Title/Abstract:~3] OR "COVID-19 vaccine"[Title/Abstract:~3] OR "COVID-19 vaccines"[Title/Abstract:~3] OR "COVID-19 vaccination"[Title/Abstract:~3] OR "COVID-19 vaccinated"[Title/Abstract:~3] OR "nCov-19 vaccine"[Title/Abstract:~3] OR "nCov-19 vaccines"[Title/Abstract:~3] OR "nCov-19 vaccination"[Title/Abstract:~3] OR "nCov-19 vaccinated"[Title/Abstract:~3] OR "2019-nCoV vaccine"[Title/Abstract:~3] OR "2019-nCoV vaccines"[Title/Abstract:~3] OR "2019-nCoV vaccination"[Title/Abstract:~3] OR "2019-nCoV vaccinated"[Title/Abstract:~3] OR "Coronavirus vaccine"[Title/Abstract:~5] OR "Coronavirus vaccines"[Title/Abstract:~3] OR "Coronavirus vaccination"[Title/Abstract:~3] OR "Coronavirus vaccinated"[Title/Abstract:~3] OR (("Pfizer-BioNTech"[Title/Abstract] OR "Comirnaty"[Title/Abstract] OR "BNT162b2"[Title/Abstract] OR "BNT162"[Title/Abstract] OR "BNT-162B2"[Title/Abstract] OR "mRNA-1273"[Title/Abstract] OR "Moderna"[Title/Abstract] OR "Spikevax"[Title/Abstract] OR "Ad26COVS1"[Title/Abstract] OR "Ad26.COV2.S"[Title/Abstract] OR "Jcovden"[Title/Abstract] OR "Janssen"[Title/Abstract] OR "ChAdOx1"[Title/Abstract] OR "ChAdOx1-S"[Title/Abstract] OR "Vaxzevria"[Title/Abstract] OR "AstraZeneca"[Title/Abstract] OR "oxford astrazeneca"[Title/Abstract] OR "AZD1222"[Title/Abstract] OR "Sinovac"[Title/Abstract] OR "CoronaVac"[Title/Abstract] OR "Sinovac-CoronaVac"[Title/Abstract] OR "Novavax"[Title/Abstract] OR "Nuvaxovid"[Title/Abstract] OR "Bimervax"[Title/Abstract] OR "HIPRA"[Title/Abstract] OR "PHH-1V"[Title/Abstract] OR ("VidPrevtyn"[All Fields] AND "Beta"[Title/Abstract]) OR "Sinopharm"[Title/Abstract] OR "Valneva"[Title/Abstract] OR "VLA2001"[Title/Abstract]) AND "vaccin*"[Title/Abstract]) OR "COVID-19 vaccines"[MeSH Terms]) AND ("Safety"[Title/Abstract] OR "harm*"[Title/Abstract] OR "adverse event*"[Title/Abstract] OR "adverse effect*"[Title/Abstract] OR "adverse drug reaction*"[Title/Abstract] OR "adverse drug event*"[Title/Abstract] OR "adverse reaction*"[Title/Abstract] OR "side effect*"[Title/Abstract] OR "complication*"[Title/Abstract] OR "undesirable effect*"[Title/Abstract] OR "unfavorable effect*"[Title/Abstract] OR "Pharmacovigilance"[Title/Abstract] OR "risk*"[Title/Abstract] OR "drug related side effects and adverse reactions"[MeSH Terms] OR "covid 19 vaccines/adverse effects"[MeSH Terms]) AND ("associate*"[Title/Abstract] OR "association*"[Title/Abstract] OR "Comparing"[Title/Abstract] OR "compare*"[Title/Abstract] OR "Comparative"[Title/Abstract] OR "Comparison"[Title/Abstract] OR "Versus"[Title/Abstract] OR "risk ratio*"[Title/Abstract] OR "RR"[Title/Abstract] OR "risk difference*"[Title/Abstract] OR "relative risk"[Title/Abstract] OR "absolute risk*"[Title/Abstract] OR "relative incidence"[Title/Abstract] OR "odds ratio*"[Title/Abstract] OR "aOR"[Title/Abstract] OR "incidence rate ratio*"[Title/Abstract] OR "IRR"[Title/Abstract] OR "IRRs"[Title/Abstract] OR "rate difference*"[Title/Abstract] OR "hazard ratio*"[Title/Abstract] OR "HR"[Title/Abstract] OR "relative rate"[Title/Abstract] OR "attributable fraction"[Title/Abstract] OR "attributable risk"[Title/Abstract] OR "attributable rate"[Title/Abstract] OR "number needed to harm"[Title/Abstract] OR "NNH"[Title/Abstract] OR "excess events"[Title/Abstract] OR "excess cases"[Title/Abstract] OR "excess number*"[Title/Abstract] OR "comparative study"[Publication Type])) NOT (("trial"[Title] AND ("randomized"[Title] OR "randomised"[Title] OR "placebo-controlled"[Title] OR "phase 1"[Title] OR "phase 2"[Title] OR "phase 3"[Title])) OR "review"[Title])) AND (2020/12/11:3000/12/12[Date - Publication] AND "english"[Language]) | | | 8,405<br><br>Oct 09, 2023 |
| **EMBASE**<br><br>(((('sars-cov-2' OR 'covid-19' OR 'ncov-19' OR '2019-ncov' OR 'coronavirus' OR 'sars-cov-2' OR 'covid-19') NEAR/3 'vaccin*'):ti,ab,kw) OR (('pfizer-biontech':ab,ti,kw OR 'comirnaty':ab,ti,kw OR 'bnt162b2':ab,ti,kw OR 'bnt162':ab,ti,kw OR 'bnt-162b2':ab,ti,kw OR 'mrna-1273':ab,ti,kw OR 'moderna':ab,ti,kw OR 'spikevax':ab,ti,kw OR 'ad26covs1':ab,ti,kw OR 'ad26.cov2.s':ab,ti,kw OR 'jcovden':ab,ti,kw OR 'janssen':ab,ti,kw OR 'chadox1':ab,ti,kw OR 'chadox1-s':ab,ti,kw OR 'vaxzevria':ab,ti,kw OR 'astrazeneca':ab,ti,kw OR 'oxford astrazeneca':ab,ti,kw OR 'azd1222':ab,ti,kw OR 'sinovac':ab,ti,kw OR 'coronavac':ab,ti,kw OR 'sinovac-coronavac':ab,ti,kw OR 'novavax':ab,ti,kw OR 'nuvaxovid':ab,ti,kw OR 'bimervax':ab,ti,kw OR 'hipra':ab,ti,kw OR 'phh-1v':ab,ti,kw OR 'sinopharm':ab,ti,kw OR 'valneva':ab,ti,kw OR 'vla2001':ab,ti,kw) AND 'vaccin*':ab,ti,kw) OR 'SARS-CoV-2 vaccine'/exp) AND ('safety':ab,ti,kw OR 'harm*':ab,ti,kw OR 'adverse event*':ab,ti,kw OR 'adverse effect*':ab,ti,kw OR 'adverse drug reaction*':ab,ti,kw OR 'adverse drug event*':ab,ti,kw OR 'adverse reaction*':ab,ti,kw OR 'side effect*':ab,ti,kw OR 'complication*':ab,ti,kw OR 'undesirable effect*':ab,ti,kw OR 'unfavorable effect*':ab,ti,kw OR 'pharmacovigilance':ab,ti,kw OR 'risk*':ab,ti,kw OR 'adverse drug reaction'/exp) AND ('associat*':ab,ti,kw OR 'comparing':ab,ti,kw OR 'compare*':ab,ti,kw OR 'comparative':ab,ti,kw OR 'comparison':ab,ti,kw OR 'versus':ab,ti,kw OR 'risk ratio*':ab,ti,kw OR 'rr':ab,ti,kw OR 'risk difference*':ab,ti,kw OR 'relative risk':ab,ti,kw OR 'absolute risk*':ab,ti,kw OR 'relative incidence':ab,ti,kw OR 'odds ratio*':ab,ti,kw OR 'aor':ab,ti,kw OR 'incidence rate ratio*':ab,ti,kw OR 'irr':ab,ti,kw OR 'irrs':ab,ti,kw OR 'rate difference*':ab,ti,kw OR 'hazard ratio*':ab,ti,kw OR 'hr':ab,ti,kw OR 'relative rate':ab,ti,kw OR 'attributable fraction':ab,ti,kw OR 'attributable risk':ab,ti,kw OR 'attributable rate':ab,ti,kw OR 'number needed to harm':ab,ti,kw OR 'nnh':ab,ti,kw OR 'excess events':ab,ti,kw OR 'excess cases':ab,ti,kw OR 'excess number*':ab,ti,kw) NOT ('clinical trial':it OR 'case reports':it OR 'clinical conference':it OR 'systematic review':it OR 'editorial':it OR 'comment':it OR 'clinical trial protocol':it OR 'review':it) AND ([article]/lim OR [article in press]/lim) AND [11-12-2020]/sd NOT [02-10-2023]/sd AND [2020-2023]/py AND [embase]/lim NOT ([embase]/lim AND [medline]/lim) | | | 2478<br><br>Oct 09, 2023 |

#### 3.4 Study selection

ASReview, an open-source machine learning algorithm for efficient systematic review will be used for records deduplication and title and abstract screening (3). The records will be screened in two rounds. In the first round, the default setting of *TF-IDF feature extraction technique, Naïve Bayes classifier, Maximum query strategy* and *Dynamic Resampling balance strategy* will be used. In the second round, the combination of *Fully connected neural network classifier, doc2vec, Maximum* and *Dynamic resampling* are chosen as the active learning model. Before the first screening round, twenty records pre-classified by the reviewer will be fed into the software to train the machine learning model. Two reviewers will independently screen the first round to compare results and resolve discrepancy in record selection. The results of the first round will be used as prior knowledge for the second round, which will be screened by one reviewer. Each screening round will be stopped after 100 consecutive records are marked as "irrelevant". Results of two rounds are combined to create a list of potentially eligible articles based on title and abstract screening. The switching-model screening strategy is recommended by the ASReview developer to enhance the coverage of relevant records (4).

After the title and abstract screening, the full-text of the potentially eligible articles will be assessed by three reviewers. Each reviewer will screen 33% of the total records and independently double check all the records of one other reviewer. Discrepancies between reviewers will be resolved by discussion among the research team.

#### 3.5 Data extraction

Data on outcomes investigated in each paper will be extracted. Then the papers investigating the three most common outcomes will be chosen for full data extraction. Each of three reviewer will extract data from 33% of the eligible papers and one reviewer will double check randomly 50% of the total eligible studies. The following information will be extracted from each paper:

- Study setting, data source and population
- Aspects of study design and statistical analysis
- Exposure, outcome and effect estimates

After extracting all outcomes and re-estimating the workload needed, the study team decided to choose to perform full data extraction for two most common outcomes – Myocarditis and Bell's Palsy only. The results of Bell's Palsy outcome will be presented in a separate paper.

#### 3.6 Risk of bias assessment

The risk of bias (RoB) of each chosen study will be independently assessed by two reviewers. Due to the observational nature of studies included in this review, the Risk of Bias in Non-randomized Studies of Interventions (ROBINS-I) tool will be used to assess the risk of bias in each study (5). The tool adopts the "target trial emulation" approach to examine bias in seven domains: confounding, selection, treatment classification, missing data, measurement and reported results. Prior to the assessment process, a target randomized trial will be specified specifically for each study, as well as the confounders and co-interventions relevant to the intervention and outcome under examination. Signalling questions within each domain of bias will be answered to provide the basis for judgement of domain-level bias as low, moderate, serious, critical or no information. The ratings across domains will be summarized to reach an overall risk of bias judgement for each outcome examined in each included study.

Several study designs, especially self-controlled designs (such as self-controlled case series (SCCS) and self-controlled risk interval (SCRI)) require certain assumptions to be met for such study to be valid. These designs also have specific features affecting risks of bias that were not covered in the ROBINS-I tool. Thus, the major validity assumptions of the SCCS and SSCRI designs will be examined prior to assessing risk of bias by the ROBINS-I tool.

The assumptions include: (1) transient or intermittent exposure; (2) an abrupt-onset event; (3) the outcome can be recurrent (in theory) for each individual, or the outcome is rare (risk of occurrence over the study period in the entire cohort is 10% or less); (4) there is variability in time or age of the event. Only studies that fulfil all validity assumptions will be proceed further to the ROBINS-I tool assessment. Furthermore, the ROBINS-I tool will be adapted with several additional or modified signaling questions to accommodate specific design features of self-controlled design.

#### 3.7 Descriptive analysis

Continuous variables will be presented as median and interquartile range. Categorical variables will be summarized with frequencies and percentages. A post-hoc analysis will be conducted to assess the consistency of results in studies where two study designs and two approaches to handle COVID-19 infection were used. Three agreement metrics will be evaluated for each pair of effect estimates(6): 1) Statistical significance agreement: whether two estimates and their confidence intervals are on the same side of the null; 2) Estimate agreement: whether the point estimate of one design/approach is within the 95% CI of the other design/approach; and 3) Standardized difference agreement: whether the absolute value of the standardized difference is < 1.96). The standardized difference is calculated as

$$Std.Diff._i = \frac{\hat{\theta}_{1i} - \hat{\theta}_{2i}}{\sqrt{se_{1i}^2 + se_{2i}^2}}$$

Where $\hat{\theta}_{1i}$, $\hat{\theta}_{2i}$ are the log of the effect estimate (HR, OR, IRR) of the design/approach 1 and 2 of study *I*, and $se_{1i}^2, se_{2i}^2$ are their associated variance.

#### 3.8 Meta-analysis

Meta-analysis of primary analyses of the study will be conducted to characterize the heterogeneity of primary effect estimates between studies. The primary analyses of the studies will be grouped by PICO question: 1) General population or population with specific conditions, 2) Vaccine brand and dose 3) unvaccinated individual/period or active comparator, and 4) outcome name. If the outcome is rare (<5% incidence), different ratio metrics (RR, OR, HR, IRR) could be combined, otherwise approximate conversions of OR and HR to RR will be applied (7). Random-effect meta-analysis will be conducted for groups with at least 3 studies. Tau-squared, $I^2$ and prediction interval will be calculated to measure degree of heterogeneity, and forest plots will be generated.

If a group includes more than one effect estimate per study or a particular study population, random-effect meta-analysis with robust variance estimation (RVE) method will be conducted to account for the dependencies between effect estimates (8). This method takes into account the dependence of the effect sizes in the dataset to provide valid inference of variance and statistical significance. RVE does not require knowledge of the exact dependence structure between effect size estimates, but use a *working model* to approximate the dependence structure. Depending on the source dependence in each PECO, the working model that best represents the dependence structure will be chosen. If the dependence arises from effect sizes estimated for non-overlapping subgroups in a study, or the same research group conducted different studies, the hierarchical effects working model will be employed. If there are effect sizes from overlapping samples, the correlated and hierarchical effects (CHE) working model will be chosen (8). The correlation between two effect sizes in the same study ($\rho$) is assumed to be constant and equal 0.7.

For studies that reported multiple effect estimates per PICO for overlapping samples (e.g. using different risk windows), decision rule will be applied to choose one single effect size per study (9).

*Table 2. Decision rules to choose independent effect sizes*

| Source of multiplicity | Decision rule |
|---|---|
| Population | Select the effect size based on the broadest population or the largest sample size<br>If the study report effect sizes for non-overlapping subgroups, the effect size for an aggregate that reassembles the full sample will be estimated |
| Intervention | Select intervention most relevant (most commonly used in the study sample).<br>If the interventions are sufficiently similar, effect sizes across interventions could be collapsed to estimate the overall effect size |
| Outcome | Select the outcome most relevant (most frequently reported across studies) |
| Risk window | Select the window most frequently reported across studies |
| Analyses | Select the analyses with the highest level of confounding/bias adjustment |

For myocarditis outcome, univariate sub-group analyses will be conducted for PICOs with at least three studies to explore the possible causes of heterogeneity in effect estimates between studies. The sub-groups included: 1) Age restriction of the study (general population, younger population only (<40 years old), or older population only (>50 years old); 2) Risk window length (within 14 days after vaccination, within 28 days after vaccination, or other lengths); 3) Study design (Cohort, SCCS/SCRI, or other designs); 4) Overall RoB (Low-moderate or serious-critical); 5) RoB of confounding domain (Low-moderate or serious-critical); 6) Outcome definition (myocarditis only, or myocarditis/pericarditis); and 7) Approach to handle COVID-19 infection in the analysis (No approach applied, approaches that measure direct effect of vaccination only, or other approaches).

Contour-enhanced funnel plots will be generated for PICO with at least 10 studies to investigate small-study effects and differentiate asymmetry that is due to non-reporting biases from that due to other factors (10-13).

#### 3.9 Examination of sources of heterogeneity by meta-regression

For groups with at least 10 studies, random-effect meta-regression with robust variance estimation will be conducted to investigate sources of heterogeneity, accounting for the dependencies between effect estimates (8). This method takes into account the dependence of the effect sizes in the dataset to provide valid inference of variance and statistical significance. RVE does not require knowledge of the exact dependence structure between effect size estimates, but use a *working model* to approximate the dependence structure. Depending on the source dependence in each PICO, the working model that best represents the dependence structure will be chosen. If the dependence arises from effect sizes estimated for non-overlapping subgroups in a study, or the same research group conducted different studies, the hierarchical effects working model will be employed. If there are effect sizes from overlapping samples, the correlated and hierarchical effects (CHE) working model will be chosen (8). The correlation between two effect sizes in the same study ($\rho$) is assumed to be constant and equal 0.7. Small sample adjustment using MBBS estimator for standard errors and p-value will be employed (14).

For myocarditis outcome, the variables in Table 3 will be investigated. The categorical variables will be dummy coded. For variables that varied both within a study and between studies (e.g. age and sex distribution, length of risk window, as studies often have subgroup/ sensitivity analysis regarding these variables), the study mean and the study-mean-centered values of those variables will be computed. All primary analyses, subgroup analyses and sensitivity analyses related to variable of interest will be included in the model to increase statistical power and to estimate their between-study and within-study effects.

The base meta-regression model includes two case-mix variables, age, and sex. Both the study means and the study-mean-centered values of these two variables will be included in the model, to estimate separately their between-study and within-study effect, respectively (15, 16). Next, the effect of setting and methodological factors will be investigated one by one, adjusted for age and sex. When estimating the effects of factors that only varied between studies (e.g. study design, RoB), only the between-study effects of age and sex covariates need to be adjusted for, thus their within-study effect (i.e. study-mean-centered values) will be omitted from the model (17). The regression coefficients (β), their associated 95% CI and the Wald-style F test p-value of each variable (indicating the overall significance of a variable) will be reported (16). Significance level will be set at 0.01, because of the inflating type-I error with small sample (15).

*Table 3. Variable of interest for meta-regression, myocarditis outcomes*

| **Characteristics** | **Definition** | **Data cleaning process** |
|---|---|---|
| Participant characteristics | | |
| **Age distribution** | Range of the median age of the study sample.<br>Three categories:<br>< 30 yo<br>30-49 yo | The median age of the study sample will be extracted.<br>If study only reported proportion of each age group, the median will be estimated (method described here). If the median lies within an age group with unknown upper or lower bound, data will be considered as missing. |

| | | |
|---|---|---|
| | 50+ | If study only reported mean ages, those values will be used (assume symmetric age distribution)<br>For subgroup analysis of age group, the mean of the lower and upper bound of the age group will be used. For age groups with unknown upper and/or lower bound, data will be considered as missing.<br>To account for the possible inaccuracy of the estimated median age, median age will be categorized into three ranges, instead of the specific continuous values.<br>For sub-group and sensitivity analysis per variables other than age (for which the median age of the sub-sample was not reported), the sub-sample will be assumed to fall into the same median age category as the main sample. This is to make use of all available information without imposing too strong assumption. |
| **Sex distribution** | Proportion of male in the study sample.<br>Five categories:<br>All female (0%)<br>Female-dominant (<45%)<br>Balance (45-55%)<br>Male-dominant (>55%)<br>All male (100%) | The proportion of male in the study sample will be extracted, then categorized into five ranges.<br>For sub-group and sensitivity analysis per variables other than sex (for which the sex distribution of the sub-sample was not reported), the sub-sample will be assumed to fall into the same sex distribution category as the main sample. |
| Design characteristics | | |
| **Study design** | 0 = Cohort<br>1 = Case-control<br>2 = Cross-sectional<br>3 = SCCS/SCRI<br>5 = Others | Categories will be collapsed to "others" if there are not enough studies per categories (<3) |
| **Length of risk window** | Categorical variables:<br>0/1-6/7 day (1$^{st}$ week)<br>0/1-20/21 day (first 3 weeks)<br>0/1-27/28 day (first 4 weeks)<br>7/8-14 day (2$^{nd}$ week)<br>14-21 day (3$^{rd}$ week)<br>21-28 day (4$^{th}$ week)<br>Others | Categories will be merged with the next category if there are not enough studies per categories (<3) |
| **How COVID-19 infection was handled** | 0 = Do not control for COVID-19<br>1 = Exclude individuals with COVID-19 before risk window | Categories will be collapsed to "others" if there are not enough studies per categories (<3) |

| | | |
|---|---|---|
| | 2 = Censor individuals with COVID-19<br>4 = Covariate in multivariable model<br>5 = Exclude individuals with COVID-19 before and during study periods<br>6 = Censor individuals with COVID-19 during study period and Covariate in multivariable model for those with COVID before study period | |
| **Outcome definition** | Two categories<br>- Myocarditis or myopericartitis<br>- Myocarditis or pericarditis | |
| **Overall RoB** | Low / Moderate / Serious / Critical / NI | Categories will be merged as low-moderate, serious-critical, NI, if there are not enough studies per category.<br>If there is no or little variation in one domain, that domain will not be included in the model. |
| **RoB: confounding** | Low / Moderate / Serious / Critical / NI | |
| **RoB: selection of participants into study** | Low / Moderate / Serious / Critical / NI | |
| **RoB: measurement of outcome** | Low / Moderate / Serious / Critical / NI | |
| **RoB: classification of intervention** | Low / Moderate / Serious / Critical / NI<br>More relevant for self-controlled studies | |
