## Supplementary file 3 for "Quality and methodological heterogeneity of COVID-19 vaccine safety studies focusing on the myocarditis safety signal: A systematic review, meta-analysis and meta-regression"

The Risk Of Bias In Non-randomized Studies – of Interventions (ROBINS-I) assessment tool

(version for SELF-CONTROLLED CASE SERIES AND SELF-CONTROLLED RISK INTERVAL studies)

**Version 21 September 2023**

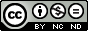

This work is licensed under a [Creative Commons Attribution-NonCommercial-NoDerivatives 4.0 International License](http://creativecommons.org/licenses/by-nc-nd/4.0/).

### ROBINS-I tool (Stage I): At protocol stage

#### Specify the review question

| Participants |
| --- |
| Experimental intervention |
| Comparator |
| Outcomes |

#### List the confounding domains relevant to all or most studies

#### List co-interventions that could be different between intervention groups and that could impact on outcomes

### ROBINS-I tool (Stage II): For each study

#### Specify a target randomized trial specific to the study

| Design | Individually randomized / Cluster randomized / Matched (e.g. cross-over) |
| --- | --- |
| Participants |  |
| Experimental intervention |  |
| Comparator |  |

#### Is your aim for this study…?

| □ | to assess the effect of *assignment to* intervention |
| --- | --- |
| □ | to assess the effect of *starting and adhering to* intervention |

#### Specify the outcome

Specify which outcome is being assessed for risk of bias (typically from among those earmarked for the Summary of Findings table). Specify whether this is a proposed benefit or harm of intervention.

#### Specify the numerical result being assessed

In case of multiple alternative analyses being presented, specify the numeric result (e.g. RR = 1.52 (95% CI 0.83 to 2.77) and/or a reference (e.g. to a table, figure or paragraph) that uniquely defines the result being assessed.

#### Preliminary consideration of confounders

Complete a row for each important confounding domain (i) listed in the review protocol; and (ii) relevant to the setting of this particular study, or which the study authors identified as potentially important.

###### “Important” confounding domains are those for which, in the context of this study, adjustment is expected to lead to a clinically important change in the estimated effect of the intervention. “Validity” refers to whether the confounding variable or variables fully measure the domain, while “reliability” refers to the precision of the measurement (more measurement error means less reliability).

| **(i) Confounding domains listed in the review protocol** | | | | |
| --- | --- | --- | --- | --- |
| Confounding domain | Measured variable(s) | Is there evidence that controlling for this variable was unnecessary?* | Is the confounding domain measured validly and reliably by this variable (or these variables)? | OPTIONAL: Is failure to adjust for this variable (alone) expected to favour the experimental intervention or the comparator? |
|  |  |  | Yes / No / No information | Favour experimental / Favour comparator / No information |

| **(ii) Additional confounding domains relevant to the setting of this particular study, or which the study authors identified as important** | | | | |
| --- | --- | --- | --- | --- |
| Confounding domain | Measured variable(s) | Is there evidence that controlling for this variable was unnecessary?* | Is the confounding domain measured validly and reliably by this variable (or these variables)? | OPTIONAL: Is failure to adjust for this variable (alone) expected to favour the experimental intervention or the comparator? |
|  |  |  | Yes / No / No information | Favour experimental / Favour comparator / No information |

* In the context of a particular study, variables can be demonstrated not to be confounders and so not included in the analysis: (a) if they are not predictive of the outcome; (b) if they are not predictive of intervention; or (c) because adjustment makes no or minimal difference to the estimated effect of the primary parameter. Note that “no statistically significant association” is not the same as “not predictive”.

#### Preliminary consideration of co-interventions

Complete a row for each important co-intervention (i) listed in the review protocol; and (ii) relevant to the setting of this particular study, or which the study authors identified as important.

###### “Important” co-interventions are those for which, in the context of this study, adjustment is expected to lead to a clinically important change in the estimated effect of the intervention.

| **(i) Co-interventions listed in the review protocol** | | |
| --- | --- | --- |
| Co-intervention | Is there evidence that controlling for this co-intervention was unnecessary (e.g. because it was not administered)? | Is presence of this co-intervention likely to favour outcomes in the experimental intervention or the comparator |
|  |  | Favour experimental / Favour comparator / No information |
|  |  | Favour experimental / Favour comparator / No information |
|  |  | Favour experimental / Favour comparator / No information |
|  |  | Favour experimental / Favour comparator / No information |

| **(ii) Additional co-interventions relevant to the setting of this particular study, or which the study authors identified as important** | | |
| --- | --- | --- |
| Co-intervention | Is there evidence that controlling for this co-intervention was unnecessary (e.g. because it was not administered)? | Is presence of this co-intervention likely to favour outcomes in the experimental intervention or the comparator |
|  |  | Favour experimental / Favour comparator / No information |
|  |  | Favour experimental / Favour comparator / No information |
|  |  | Favour experimental / Favour comparator / No information |
|  |  | Favour experimental / Favour comparator / No information |

#### Risk of bias assessment

Responses underlined in green are potential markers for low risk of bias, and responses in red are potential markers for a risk of bias. Where questions relate only to sign posts to other questions, no formatting is used.

|  | **Signalling questions** | **Description** | **Response options** |
| --- | --- | --- | --- |
| **Bias due to confounding** | | | |
|  | 1.1 Is there potential for confounding of the effect of intervention in this study?  **If N/PN to 1.1:** the study can be considered to be at low risk of bias due to confounding and no further signalling questions need be considered |  | Y / PY / PN / N |
|  | **If Y/PY to 1.1**: determine whether there is a need to assess time-varying confounding: |  |  |
|  | 1.2. Was the analysis based on splitting participants’ follow up time according to intervention received?  **If N/PN**, answer questions relating to baseline confounding (1.4 to 1.6)  **If Y/PY**, go to question 1.3. |  | NA / Y / PY / PN / N / NI |
|  | 1.3. Were intervention discontinuations or switches likely to be related to factors that are prognostic for the outcome?  **If N/PN**, answer questions relating to baseline confounding (1.4 to 1.6)  **If Y/PY**, answer questions relating to both baseline and time-varying confounding (1.7 and 1.8) |  | NA / Y / PY / PN / N / NI |

|  | **Questions relating to baseline confounding only** | | |
| --- | --- | --- | --- |
|  | 1.4. Did the authors use an appropriate analysis method that controlled for all the important confounding domains? ** |  | NA / Y / PY / PN / N / NI |
|  | 1.5. **If Y/PY to 1.4**: Were confounding domains that were controlled for measured validly and reliably by the variables available in this study? |  | NA / Y / PY / PN / N / NI |
|  | 1.6. Did the authors control for any post-intervention variables that could have been affected by the intervention? |  | NA / Y / PY / PN / N / NI |
|  | **Questions relating to baseline and time-varying confounding** | |  |
|  | 1.7. Did the authors use an appropriate analysis method that controlled for all the important confounding domains and for time-varying confounding? |  | NA / Y / PY / PN / N / NI |
|  | 1.8. **If Y/PY to 1.7**: Were confounding domains that were controlled for measured validly and reliably by the variables available in this study? |  | NA / Y / PY / PN / N / NI |
|  | **Risk of bias judgement** |  | Low / Moderate / Serious / Critical / NI |
|  | Optional: What is the predicted direction of bias due to confounding? |  | Favours experimental / Favours comparator / Unpredictable |

*** For self-controlled study, time-invariant confounders are automatically controlled for, thus only time-varying confounders are in consideration.*

| **Bias in selection of participants into the study** | | | |
| --- | --- | --- | --- |
|  | 2.1. Was selection of participants into the study (or into the analysis) based on participant characteristics observed after the start of intervention?*  **If N/PN to 2.1:** go to 2.4 |  | Y / PY / PN / N / NI |
|  | 2.2. **If Y/PY to 2.1**: Were the post-intervention variables that influenced selection likely to be associated with intervention?  2.3 **If Y/PY to 2.2**: Were the post-intervention variables that influenced selection likely to be influenced by the outcome or a cause of the outcome? |  | NA / Y / PY / PN / N / NI  NA / Y / PY / PN / N / NI |
|  | 2.4. Do start of follow-up and start of intervention coincide for most participants?** |  | Y / PY / PN / N / NI |
|  | 2.5. **If Y/PY to 2.2 and 2.3, or N/PN to 2.4**: Were adjustment techniques used that are likely to correct for the presence of selection biases? |  | NA / Y / PY / PN / N / NI |
|  | **Risk of bias judgement** |  | Low / Moderate / Serious / Critical / NI |
|  | Optional: What is the predicted direction of bias due to selection of participants into the study? |  | Favours experimental / Favours comparator / Towards null /Away from null / Unpredictable |

* *In SCCS and SCRI design, selection of study participants is based on the occurrence of outcome, and this is not considered as bias.*

*** This question is not applicable for self-controlled design*

| **Bias in classification of interventions** | | | |
| --- | --- | --- | --- |
|  | 3.1 Were intervention groups clearly defined? |  | Y / PY / PN / N / NI |
|  | 3.2 Was the information used to define intervention groups recorded at the start of the intervention? |  | Y / PY / PN / N / NI |
|  | 3.3 Could classification of intervention status have been affected by knowledge of the outcome or risk of the outcome? |  | Y / PY / PN / N / NI |
|  | **For self-controlled design: answer additional questions 3.4 – 3.9:** | | |
|  | 3.4. Is the probability of future exposure affected by a previous event? |  | Y / PY / PN / N / NI |
|  | 3.5. **If Y / PY to 3.4:** Is the design adapted by excluding pre-exposure period from the control window, or use a proper modified SCCS model? |  | Y / PY / PN / N / NI |
|  | 3.6 Is the future exposure **always** censored by the event (e.g. death as outcome)? |  | Y / PY / PN / N / NI |
|  | 3.7 **If Y / PY to 3.6:** Is the design adapted by beginning observation period at exposure? |  | Y / PY / PN / N / NI |
|  | 3.8 Does the event affect short-term mortality ? |  | Y / PY / PN / N / NI |
|  | 3.9 If **Y/PY to 3.8:** Is the design adapted:  (1) Involving the time interval between the event and end of the actual observation period  (2) Sensitivity analysis: all cases vs. excluding those who died.  (3) Correct bias by fitting an extension that involves modelling post-event survival time |  | Y / PY / PN / N / NI |
|  | **Risk of bias judgement** |  | Low / Moderate / Serious / Critical / NI |
|  | Optional: What is the predicted direction of bias due to classification of interventions? |  | Favours experimental / Favours comparator / Towards null /Away from null / Unpredictable |

| **Bias due to deviations from intended interventions** | | | |
| --- | --- | --- | --- |
|  | **If your aim for this study is to assess the effect of assignment to intervention, answer questions 4.1 and 4.2** | |  |
|  | 4.1. Were there deviations from the intended intervention beyond what would be expected in usual practice? |  | Y / PY / PN / N / NI |
|  | 4.2. **If Y/PY to 4.1**: Were these deviations from intended intervention unbalanced between groups *and* likely to have affected the outcome? |  | NA / Y / PY / PN / N / NI |
|  | **If your aim for this study is to assess the effect of starting and adhering to intervention, answer questions 4.3 to 4.6** | |  |
|  | 4.3. Were important co-interventions balanced across intervention groups? |  | Y / PY / PN / N / NI |
|  | 4.4. Was the intervention implemented successfully for most participants? |  | Y / PY / PN / N / NI |
|  | 4.5. Did study participants adhere to the assigned intervention regimen? |  | Y / PY / PN / N / NI |
|  | 4.6. **If N/PN to 4.3, 4.4 or 4.5**: Was an appropriate analysis used to estimate the effect of starting and adhering to the intervention? |  | NA / Y / PY / PN / N / NI |
|  | **Risk of bias judgement** |  | Low / Moderate / Serious / Critical / NI |
|  | Optional: What is the predicted direction of bias due to deviations from the intended interventions? |  | Favours experimental / Favours comparator / Towards null /Away from null / Unpredictable |

| **Bias due to missing data** | | | |
| --- | --- | --- | --- |
|  | 5.1 Were outcome data available for all, or nearly all, participants?* |  | Y / PY / PN / N / NI |
|  | 5.2 Were participants excluded due to missing data on intervention status? |  | Y / PY / PN / N / NI |
|  | 5.3 Were participants excluded due to missing data on other variables needed for the analysis? |  | Y / PY / PN / N / NI |
|  | 5.4 **If PN/N to 5.1, or Y/PY to 5.2 or 5.3**: Are the proportion of participants and reasons for missing data similar across interventions? |  | NA / Y / PY / PN / N / NI |
|  | 5.5 **If PN/N to 5.1, or Y/PY to 5.2 or 5.3**: Is there evidence that results were robust to the presence of missing data? |  | NA / Y / PY / PN / N / NI |
|  | **Risk of bias judgement** |  | Low / Moderate / Serious / Critical / NI |
|  | Optional: What is the predicted direction of bias due to missing data? |  | Favours experimental / Favours comparator / Towards null /Away from null / Unpredictable |

** For self-controlled design, “participants” should be understood as “study population”, as the selection of participants are based on availability of outcome.*

| **Bias in measurement of outcomes** | | | |
| --- | --- | --- | --- |
|  | 6.1 Could the outcome measure have been influenced by knowledge of the intervention received? |  | Y / PY / PN / N / NI |
|  | 6.2 Were outcome assessors aware of the intervention received by study participants? |  | Y / PY / PN / N / NI |
|  | 6.3 Were the methods of outcome assessment comparable across intervention groups? |  | Y / PY / PN / N / NI |
|  | 6.4 Were any systematic errors in measurement of the outcome related to intervention received? |  | Y / PY / PN / N / NI |
|  | **For self-controlled design: answer additional questions 6.5 – 6.7:** | | |
|  | 6.5 Are two consecutive event independent if they are recurrent? |  | Y / PY / PN / N / NI |
|  | 6.6 **If** **N/PN to 6.5**: is the study design adapted to allows a first event to increase the future event risk? |  | Y / PY / PN / N / NI/ NA |
|  | 6.7. Is the risk period appropriately chosen based on prior hypotheses, previous studies, and presumed biological mechanisms? |  | Y / PY / PN / N / NI/ NA |
|  | **Risk of bias judgement** |  | Low / Moderate / Serious / Critical / NI |
|  | Optional: What is the predicted direction of bias due to measurement of outcomes? |  | Favours experimental / Favours comparator / Towards null /Away from null / Unpredictable |

| **Bias in selection of the reported result** | | | |
| --- | --- | --- | --- |
|  | Is the reported effect estimate likely to be selected, on the basis of the results, from... |  |  |
|  | 7.1. ... multiple outcome *measurements* within the outcome domain? |  | Y / PY / PN / N / NI |
|  | 7.2 ... multiple *analyses* of the intervention-outcome relationship? |  | Y / PY / PN / N / NI |
|  | 7.3 ... different *subgroups*? |  | Y / PY / PN / N / NI |
|  | **Risk of bias judgement** |  | Low / Moderate / Serious / Critical / NI |
|  | Optional: What is the predicted direction of bias due to selection of the reported result? |  | Favours experimental / Favours comparator / Towards null /Away from null / Unpredictable |

| **Overall bias** | | | |
| --- | --- | --- | --- |
|  | **Risk of bias judgement** |  | Low / Moderate / Serious / Critical / NI |
|  | Optional: What is the overall predicted direction of bias for this outcome? |  | Favours experimental / Favours comparator / Towards null /Away from null / Unpredictable |

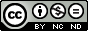

This work is licensed under a [Creative Commons Attribution-NonCommercial-NoDerivatives 4.0 International License](http://creativecommons.org/licenses/by-nc-nd/4.0/).
